# Human mobility impacts the transmission of common respiratory viruses: A modeling study of the Seattle metropolitan area

**DOI:** 10.1101/2023.10.31.23297868

**Authors:** Amanda C. Perofsky, Chelsea Hansen, Roy Burstein, Shanda Boyle, Robin Prentice, Cooper Marshall, David Reinhart, Ben Capodanno, Melissa Truong, Kristen Schwabe-Fry, Kayla Kuchta, Brian Pfau, Zack Acker, Jover Lee, Thomas R. Sibley, Evan McDermot, Leslie Rodriguez-Salas, Jeremy Stone, Luis Gamboa, Peter D. Han, Amanda Adler, Alpana Waghmare, Michael L. Jackson, Mike Famulare, Jay Shendure, Trevor Bedford, Helen Y. Chu, Janet A. Englund, Lea M. Starita, Cécile Viboud

## Abstract

Many studies have used mobile device location data to model SARS-CoV-2 dynamics, yet relationships between mobility behavior and endemic respiratory pathogens are less understood. We studied the impacts of human mobility on the transmission of SARS-CoV-2 and 16 endemic viruses in Seattle over a 4-year period, 2018-2022. Before 2020, school-related foot traffic and large-scale population movements preceded seasonal outbreaks of endemic viruses. Pathogen circulation dropped substantially after the initiation of stay-at-home orders in March 2020. During this period, mobility was a positive, leading indicator of transmission of all endemic viruses and lagged SARS-CoV-2 activity. Mobility was briefly predictive of SARS-CoV-2 transmission when restrictions relaxed in summer 2020 but associations weakened in subsequent waves. The rebound of endemic viruses was heterogeneously timed but exhibited stronger relationships with mobility than SARS-CoV-2. Mobility is most predictive of respiratory virus transmission during periods of dramatic behavioral change, and, to a lesser extent, at the beginning of epidemic waves.

**Teaser:** Human mobility patterns predict the transmission dynamics of common respiratory viruses in pre- and post-pandemic years.

## Introduction

In early 2020, there was widespread adoption of public health measures to slow the spread of severe acute respiratory syndrome coronavirus 2 (SARS-CoV-2). A variety of non-pharmaceutical interventions (NPIs) were implemented in most countries to reduce contacts between infected and susceptible individuals, including shelter-in-place or stay-at-home orders, gathering restrictions, school and business closures, and travel bans. These interventions were effective at reducing not only SARS-CoV-2 transmission but also the spread of other directly transmitted respiratory pathogens (*1–9*). Many endemic respiratory viruses did not return to widespread circulation until the end of 2020 or 2021 (*3, 8–10*), coinciding with the gradual lifting of social distancing measures and mask mandates.

During the COVID-19 pandemic, aggregated location data from mobile phones became an important source of information on changes in population-level movements and were used to model SARS-CoV-2 dynamics and assess the effectiveness of NPIs on SARS-CoV-2 transmission (*11–13*). However, few studies have explored relationships between human mobility and the dynamics of endemic respiratory pathogens during the pandemic. Here we define “endemic” pathogens as those that have regular periodic cycles and stable rates of infection in outbreak periods. Due to lack of circulation of endemic respiratory viruses in the first years of the pandemic, population susceptibility to these pathogens is expected to have increased, leading to earlier, larger, or more severe epidemics a few months later (*14, 15*). Understanding the influence of mobility patterns on the dynamics of endemic pathogens is important for predictive purposes, especially as perturbed circulation can lead to overlapping epidemics of different pathogens and in turn put extreme pressure on the healthcare system (e.g., the US “tripledemic” during winter 2022-2023)(*16*).

Here, we leverage fine-grained respiratory surveillance data and mobile device location data to explore links between population behavior and the transmission of 17 common respiratory viruses in the greater Seattle, Washington region over a 4-year period, 2018 – 2022. These viruses include SARS-CoV-2, influenza viruses (A/H3N2, A/H1N1, and B), respiratory syncytial viruses (RSV A and B), seasonal coronaviruses (hCoV 229E, OC43, HKU1, and NL63), human metapneumovirus (hMPV), human parainfluenza viruses (hPIV 1, 2, 3, and 4), human rhinovirus (hRV), and adenovirus (AdV).

## Results

### Study overview

We use detailed individual-level surveillance data from the Seattle Flu Study (SFS), which launched in the Fall of 2018 to improve detection and control of epidemics and pandemics (*17*). SFS carried out intensive hospital and community-based surveillance with systematic molecular testing of nasal swabs for up to 26 respiratory pathogens (*17*) (Table S1). Our study spans November 19, 2018, to June 30, 2022, during which respiratory specimens were collected from individuals with and without respiratory illness across a variety of sites throughout the Seattle metropolitan region, as previously described (*17–23*). In total, 138,060 respiratory specimens were screened for the presence of 24 or 26 pathogens (Table S1), and we retained 80,846 specimens after limiting our analysis to symptomatic individuals and discarding samples with missing metadata or from multiple testing (Table 1, Table S2). 25.3% (N = 20,640) of samples were collected in hospitals, and 74.5% (N = 60,206) were collected through community-based testing, including outpatient clinics, kiosks stationed in high foot traffic areas (*17*), swab-and-send at-home testing programs (*20, 22*), and King County COVID-19 drive through testing sites (Table 1, Table S2). The majority of hospital residuals were collected from younger age groups, while most community-based samples were collected from adults (Table 1, Figure S1, Table S2).

**Table 1.** Participant characteristics. Home residence, sex, and age distributions for individuals contributing respiratory specimens to different Seattle Flu Study (SFS) surveillance arms, including hospitals, SFS community testing (e.g., kiosks, swab-and-send at-home testing, outpatient clinics), and Public Health – Seattle & King County (PHSKC) COVID-19 drive through testing sites.

|  |  | Site Category |  |  |
| --- | --- | --- | --- | --- |
| Variable | Overall,<br>N = 80,846<sup>1</sup> | Hospital<br>residuals,<br>N = 20,640<sup>1</sup> | SFS community<br>surveillance,<br>N = 52,272<sup>1</sup> | PHSKC COVID-19<br>drive thru sites,<br>N = 7,934<sup>1</sup> |
| **Home residence** |  |  |  |  |
| North King County | 47,385 (59%) | 10,456 (51%) | 31,338 (60%) | 5,591 (70%) |
| South King County | 17,955 (22%) | 4,430 (21%) | 11,846 (23%) | 1,679 (21%) |
| Puget Sound, non-King County<sup>2</sup> | 15,506 (19%) | 5,754 (28%) | 9,088 (17%) | 664 (8.4%) |
| **Sex** |  |  |  |  |
| Female | 44,670 (56%) | 9,399 (46%) | 30,887 (59%) | 4,384 (56%) |
| Male | 35,771 (44%) | 11,238 (54%) | 21,071 (41%) | 3,462 (44%) |
| Other | 44 (<0.1%) | 0 (0%) | 44 (<0.1%) | 0 (0%) |
| (Missing) | 361 | 3 | 270 | 88 |
| **Mean age** | 31 (21) | 15 (20) | 35 (18) | 47 (18) |
| **Age group** |  |  |  |  |
| <1 | 3,455 (4.3%) | 2,986 (14%) | 469 (0.9%) | 0 (0%) |
| 1-4 | 9,834 (12%) | 6,363 (31%) | 3,341 (6.4%) | 130 (1.6%) |
| 5-17 | 10,690 (13%) | 5,823 (28%) | 4,585 (8.8%) | 282 (3.6%) |
| 18-49 | 40,297 (50%) | 3,307 (16%) | 33,019 (63%) | 3,971 (50%) |
| 50-64 | 10,897 (13%) | 1,209 (5.9%) | 7,515 (14%) | 2,173 (27%) |
| ≥65 | 5,673 (7.0%) | 952 (4.6%) | 3,343 (6.4%) | 1,378 (17%) |
| **Broad age group** |  |  |  |  |
| <5 | 13,289 (16%) | 9,349 (45%) | 3,810 (7.3%) | 130 (1.6%) |
| ≥5 | 67,557 (84%) | 11,291 (55%) | 48,462 (93%) | 7,804 (98%) |
| <sup>1</sup>n (%) |  |  |  |  |
| <sup>2</sup>Pierce, Snohomish, Kitsap, San Juan, Whatcom, Skagit, Island, Clallam, Jefferson, Mason, and Thurston counties. |  |  |  |  |

Over the course of the four-year study, 40.6% (N = 32,841) of specimens tested positive for at least one respiratory pathogen (including SARS-CoV-2), 32.4% (N = 26,182) were positive for at least one endemic respiratory pathogen, and 9.1% (N = 7,374) were positive for more than one pathogen. Prior to the start of Washington’s COVID-19 restrictions in March 2020, the most prevalent pathogens among positive samples were influenza A/H1N1 virus (17.9%), followed by hRV (15.4%), influenza A/H3N2 virus (13.9%), influenza B virus (12.2%), and RSV A (9.7%) (Figure S2). After March 2020, the most prevalent pathogens were SARS-CoV-2 (39.5%), hRV (35.4%), and AdV (5.1%) (Figure S2).

We reconstructed daily incidences for SARS-CoV-2 and each endemic pathogen, adjusting for testing volume over time, age, clinical setting, and local syndromic respiratory illness rates (Figure S3). Although SFS tested respiratory specimens for up to 26 pathogens, we limited our analysis to 17 viruses with sufficient sampling (≥ 400 positive samples during 2018-2022), including SARS-CoV-2, influenza A/H1N1, A/H3N2, and B viruses, RSV A and B, hCoV 229E, OC43, HKU1, and NL63, hPIV 1, 2, 3, and 4, hMPV, hRV, and AdV (Figure 1, Figure S3). Due to laboratory assay limitations (Table S1), we grouped epidemiologically distinct strains into one incidence time series each for hCoV 229E and hCoV OC43 (hereon hCoV 229E + OC43), hCoV HKU1 and hCoV NL63 (hCoV HKU1 + NL63), hPIV 1 and hPIV 2 (hPIV 1 + 2), hPIV 3 and hPIV 4 (hPIV 3 + 4), hRV, and AdV. hPIV 3 likely comprises most of hPIV 3 + 4 incidence because hPIV 4 infections are detected infrequently and tend to be mild or asymptomatic (*24*).

**Figure 1.**
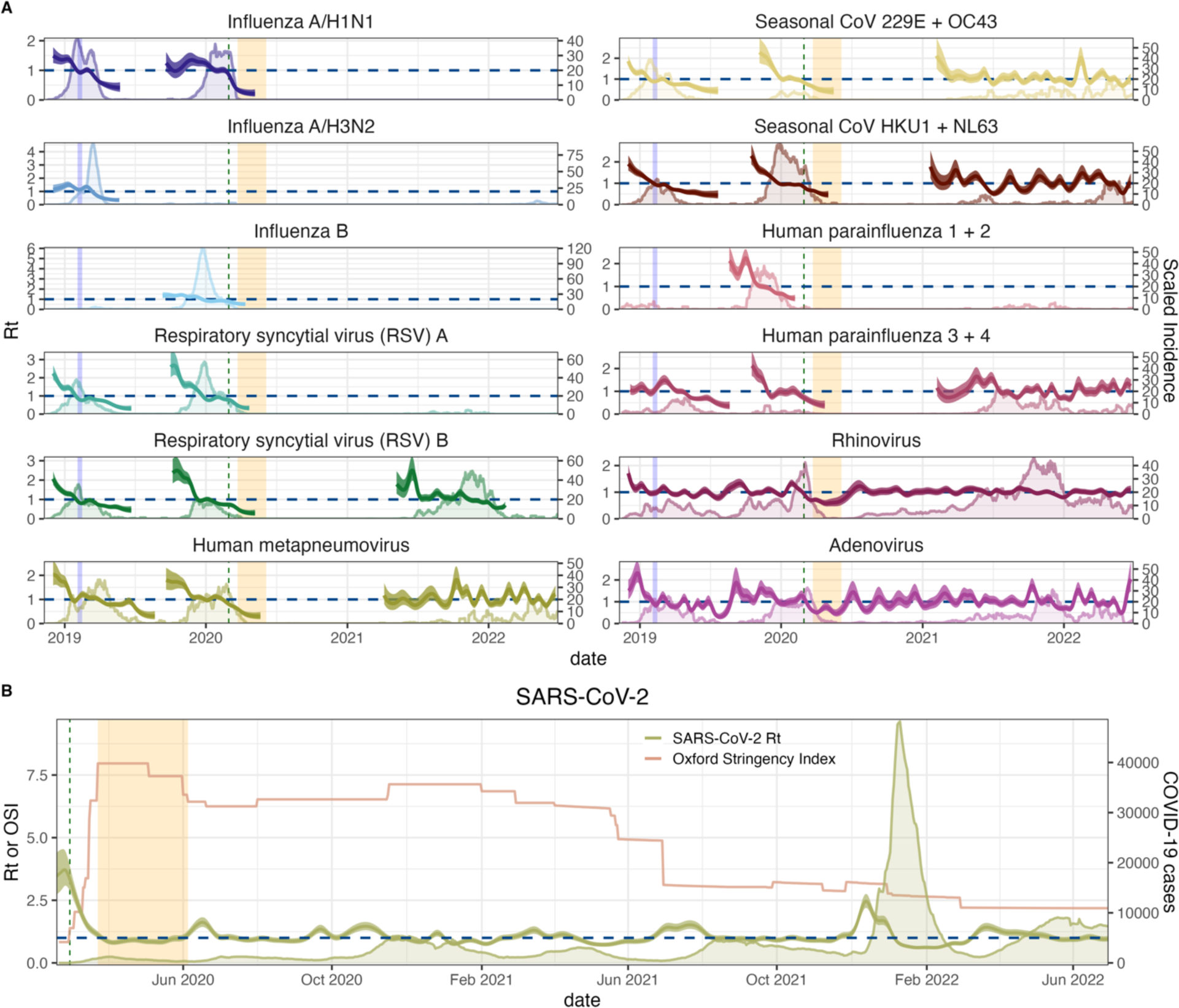
Daily incidence and transmissibility of endemic respiratory viruses and SARS-CoV-2 in the greater Seattle region. **A.** Daily time-varying effective reproduction numbers (Rt, thick lines, left y-axis) and reconstructed incidences of endemic respiratory viruses (thin lines, right y-axis) during November 2018 – June 2022. The vertical blue shaded panel indicates the timing of a major snowstorm in Seattle (February 3-15, 2019), the vertical dashed line indicates the date of Washington’s State of Emergency declaration (February 29, 2020), and the vertical orange shaded panel indicates Seattle’s stay-at-home period (March 23 – June 5, 2020). **B.** Daily time-varying effective reproduction numbers of SARS-CoV-2 (Rt, thick green line, left y-axis), King County COVID-19 case counts (thin green line, right y-axis), and the stringency of non-pharmaceutical interventions in Washington, measured by the Oxford Stringency Index (thin orange line, left y-axis), during January 2020 – June 2022.

Based on reconstructed incidences, we used semi-mechanistic epidemiological models to measure the time-varying intensity of transmission via the daily effective reproduction number (Rt)(*25, 26*) (Figure 1). To generate Rt based on dates of infection, we convolved over uncertain incubation periods and reporting delay distributions (i.e., delays from symptom onset to testing), wherein delays were informed by our individual-level surveillance data. We used aggregated mobile device location data from SafeGraph and Meta Data for Good to assess the effects of population-level movements on citywide respiratory virus dynamics in pre- and post-pandemic years (Figures 2-3). During the pandemic period, we also considered the effects of non-mobility behavioral indicators, including the stringency of Washington’s government response to COVID-19, measured by the Oxford Stringency Index (*27*) (Figure 1), and the proportion of individuals masking in public (*28*) (Figure 2).

**Figure 2.**
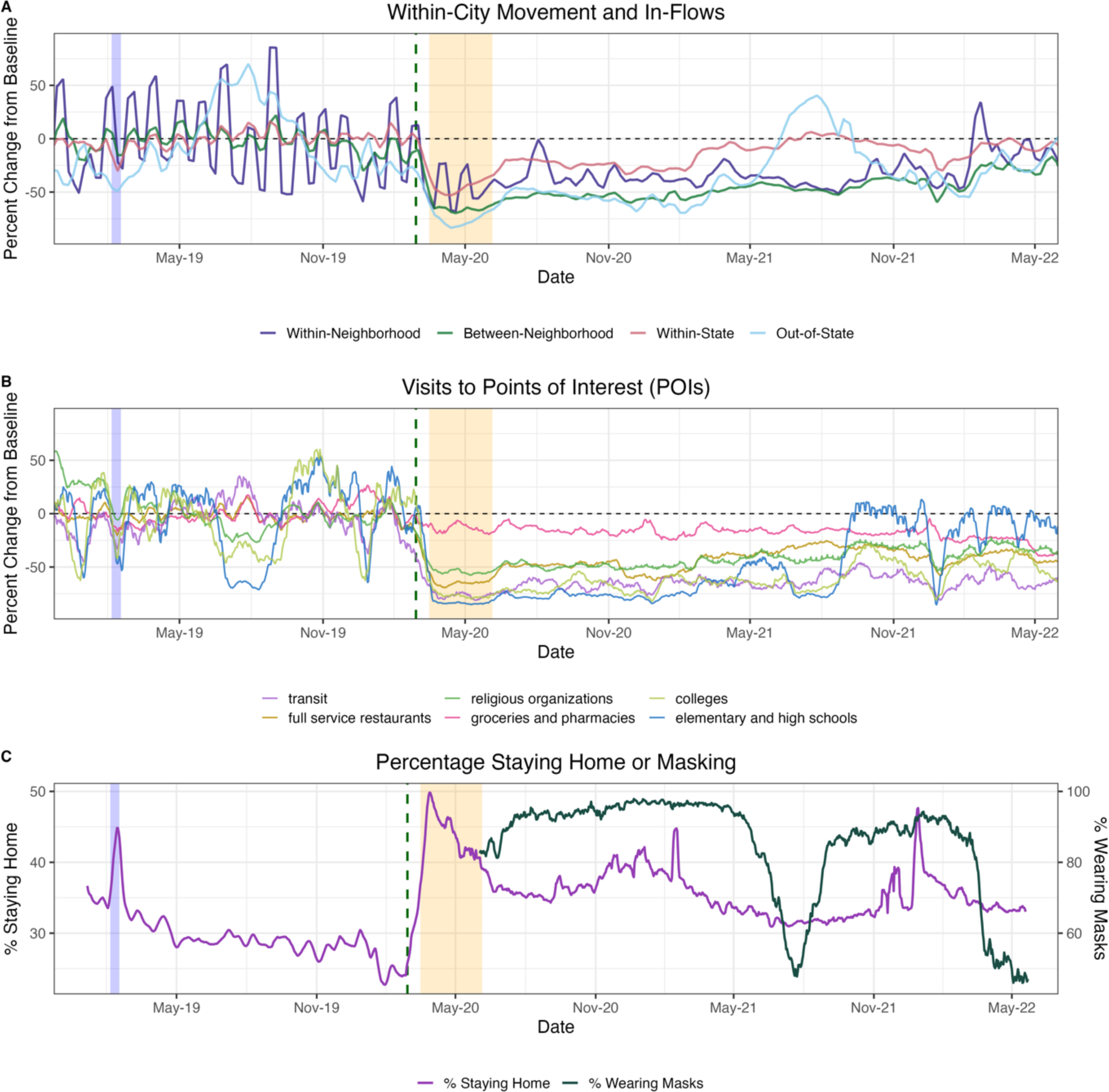
Mobility and behavior trends in the greater Seattle region based on aggregated mobile device location data, November 2018 – June 2022. In each panel, the vertical blue shaded panel indicates the timing of a major snowstorm in Seattle (February 3 – 15, 2019), the vertical dashed line indicates the date of the Washington’s State of Emergency declaration (February 29, 2020), and the vertical orange shaded panel indicates Seattle’s stay-at-home period (March 23 – June 5, 2020). **A.** The percent change from baseline for large-scale population movements: within-neighborhood movement of King County residents (purple), between-neighborhood movement of King County residents (dark green), inflow of visitors from other WA counties (red), and inflow of out-of-state visitors (light blue). **B.** The percent change from baseline in foot traffic to different categories of points of interest (POIs): transit stations (purple), religious organizations (dark green), colleges and universities (light green), full-service restaurants (dark yellow), groceries and pharmacies (pink), and elementary and high schools (blue). **C.** The percentage of devices staying completely at home (purple, left y-axis) and the percentage of individuals masking in public (dark green, right y-axis) in King County.

**Figure 3.**
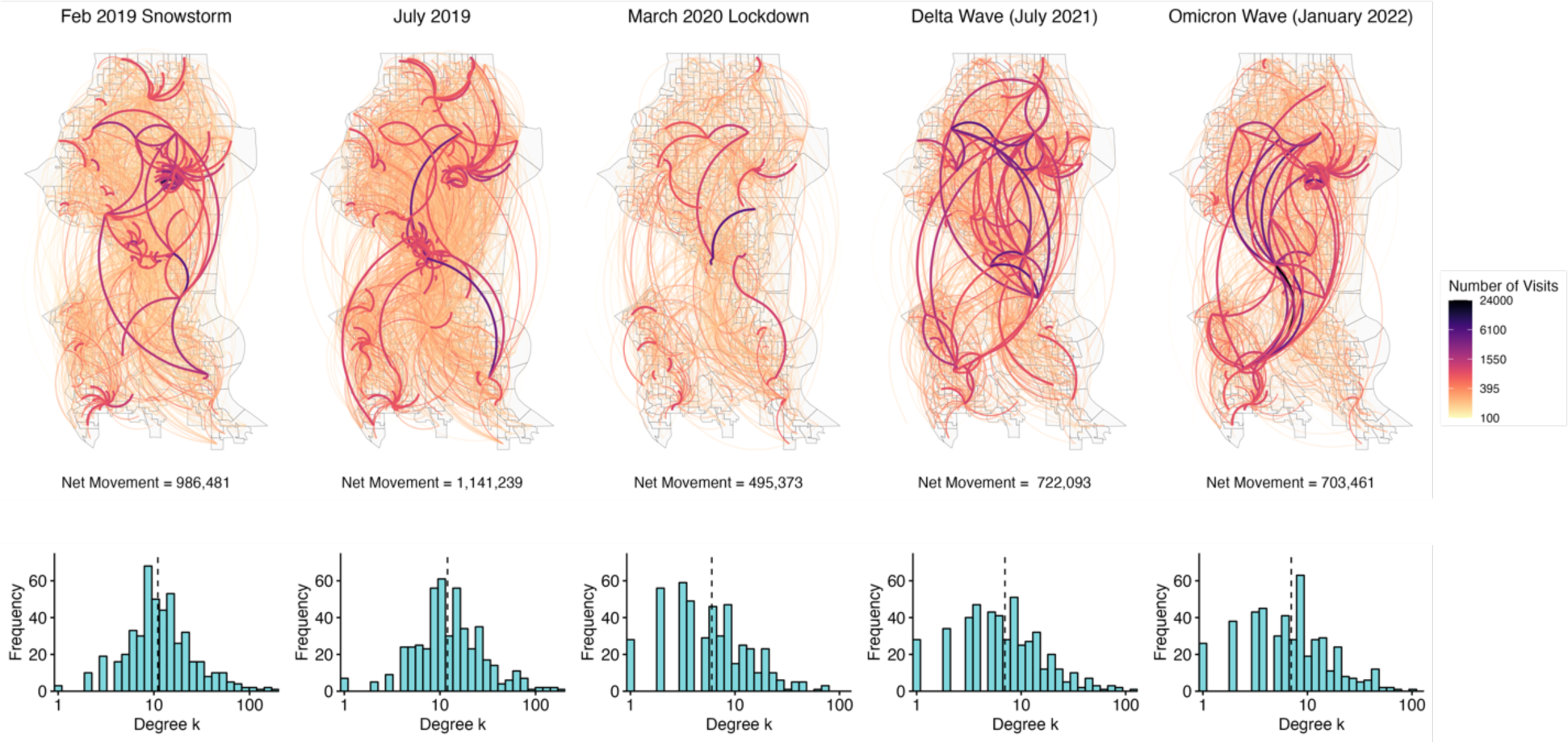
Undirected network of mobile device movement between neighborhoods (census block groups, CBGs) in Seattle, Washington, during key epidemiological time points. Time points include the week during a major snowstorm in February 2019, a week in July 2019 to show baseline movement, the beginning of stay-at-home orders in March 2020, a week during the Delta wave in July 2021, and a week during the Omicron BA.1 wave in January 2022. **Top:** Weekly visitors to points of interest (POI) are aggregated by visitor home CBG and POI CBG. Network edges (lines) are shaded according to the number of unique visitors between each pair of CBGs within a particular week, with thicker, darker edges indicating a greater number of visitors. **Bottom:** Histograms showing the frequency of degree k values for Seattle neighborhoods (i.e., the integer number of other neighborhoods each individual neighborhood is connected to) at each time point. The vertical dashed line overlaying each histogram indicates the median degree among Seattle neighborhoods.

### Declines in mobility correlate with reduced respiratory virus circulation during a major snowstorm in February 2019

Most endemic viruses in our study, including influenza A viruses, RSV, hCoV, hPIV 3+4, hMPV, hRV, and AdV, circulated during the 2018-2019 winter season. This season was atypical in that Seattle experienced unusually high snowfall during February 2019, prompting widespread school and workplace closures and reduced regional travel from February 3 to February 15, 2019. The mobility categories most impacted by the snowstorm included foot traffic to elementary and high schools, colleges, and transit stations (> 75% declines below baseline), the inflow of out-of-state visitors (> 50% declines below baseline), and within- and between-neighborhood movement (29% and 15% declines below baseline) (Figure 2, Figure S4). As previously described (*21*), this city-wide shutdown led to a conspicuous dip in incidence for several pathogens (Figure S5).

To measure the overall impact of the snowstorm on virus circulation, we compared pathogen specific Rt values during the two weeks before and after the start of heavy snowfall on February 3, 2019 (Table 2). RSV and AdV were the pathogens most affected by weather-related disruptions (37-40% declines), followed by influenza viruses and hCoV (10-20% declines, Table 2). Influenza A/H3N2 virus, hPIV 3 + 4, hMPV, hRV, and AdV rebounded after schools and workplaces reopened, and their epidemics subsequently peaked from mid-March to early April 2019 (Figure S5).

**Table 2.** Changes in transmissibility (time-varying effective reproduction numbers, Rt) during the two weeks before and after two events: a major snowstorm in February 2019 and the initiation of COVID-19 social distancing measures in March 2020. We compared Rt values before and after each event using t-tests for the ratio of two means. Fieller’s theorem was used to calculate the 95% confidence intervals of changes in Rt.

| Pathogen | Major snowstorm, February 2019 |  | Early COVID-19 restrictions, March 2020 |  |
| --- | --- | --- | --- | --- |
|  | Change in Rt | P-value | Change in Rt | P-value |
| Influenza A/H3N2 | -12 [-17, -8] % | 0.00002 | Not circulating |  |
| Influenza A/H1N1 | -19 [-25, -14] % | 0.00001 | -42 [-59, -29] % | $< 2 \times 10^{-16}$ |
| Influenza B | Not circulating | | -14 [-19, -10] % | $< 2 \times 10^{-16}$ |
| RSV A | -40 [-50, -30] % | $< 2 \times 10^{-16}$ | -9 [-14, -4] % | 0.0006 |
| RSV B | -37 [-45, -28] % | $< 2 \times 10^{-16}$ | -4 [-5, -2] % | 0.0003 |
| hMPV | 20 [16, 23] % | $< 2 \times 10^{-16}$ | -20 [-26, -15] % | $< 2 \times 10^{-16}$ |
| hPIV 1 + 2 | Not circulating |  | Not circulating |  |
| hPIV 3 + 4 | 14 [9, 18] % | 0.00004 | -22 [-28, -16] % | $< 2 \times 10^{-16}$ |
| hCoV 229E + OC43 | -10 [-15, -5] % | 0.001 | -20 [-25, -16] % | $< 2 \times 10^{-16}$ |
| hCoV HKU1 + NL63 | -18 [-22, -14] % | $< 2 \times 10^{-16}$ | -25 [-31, -20] % | $< 2 \times 10^{-16}$ |
| hRV | 2 [-0.03, 3] % | $> 0.05$ | -29 [-37, -22] % | $< 2 \times 10^{-16}$ |
| AdV | -39 [-47, -31] % | $< 2 \times 10^{-16}$ | -33 [-45, -22] % | $< 2 \times 10^{-16}$ |
| SARS-CoV-2 | Not circulating | | -37 [-51, -25] % | $< 2 \times 10^{-16}$ |

During February 2019, reductions in mobility preceded or coincided with declines in pathogen transmission, though the strength of correlations varied across pathogens (Figure S6-S7). Among pathogens with the most substantial declines, drops in RSV Rt coincided most closely with reductions in within-city connectivity and foot traffic to schools, child daycare centers, and religious services (February 2019 mean cross-correlation coefficients, r > 0.86; all reported correlations are statistically significant), while AdV Rt was moderate-to-strongly correlated with most mobility indicators, in particular visitor inflow, within-neighborhood movement, and visits to schools, child daycares, colleges, and religious services (r > 0.94) (Figure S6-S7). For pathogens that did not experience declines in transmission (hPIV 3 + 4, hMPV, hRV), Rt had negative or non-significant associations with mobility during the snowstorm (Figure S6-S7).

### Relationships between mobility and pathogen transmission during the 2019-2020 winter season (pre-pandemic)

The 2019-2020 virus respiratory season was a relatively typical season in Seattle with heightened activity of many common respiratory viruses (Figure 1). During Fall 2019, visits to child daycares, schools, colleges, and religious organizations preceded or coincided with initial increases in influenza viruses, RSV, hMPV, hCoV, and hPIV 3 + 4 (moving window cross-correlation coefficients, r range: 0.53 – 0.97; all reported correlations are statistically significant; Figure 4, Figure S8). The transmission rates of RSV, hCoV, and hPIV 3 + 4 were also positively correlated with the percentage of devices leaving home (r: 0.68 – 0.81) (Figure 4, Figure S8). Increases in hPIV 1 + 2 and AdV Rt coincided with visits to child daycares (both viruses, r: 0.63 – 0.88), between-neighborhood movement (AdV, r: 0.6 – 0.8), or out-of-state inflow (hPIV 1 + 2, r: 0.7 – 0.77), while hRV dynamics were not strongly tied to population mobility (Figure 4, Figure S8). For most pathogens, the strongest relationships between transmission and mobility occurred at the beginning of the season in early autumn (Figure 4, Figure S8).

**Figure 4.**
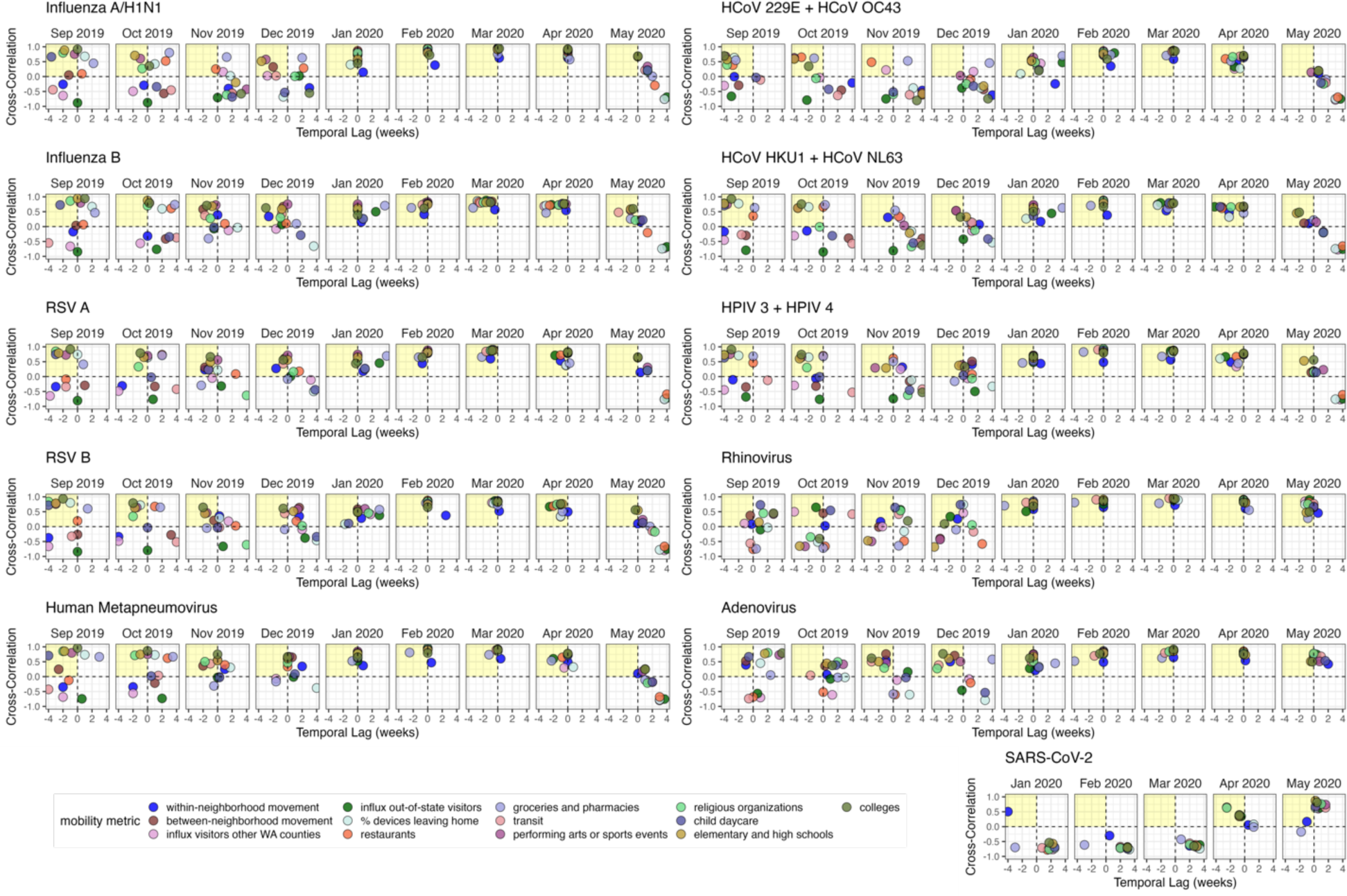
Time series cross-correlations and optimal lags between respiratory virus transmissibility (time-varying effective reproduction numbers, Rt) and cell phone mobility in the greater Seattle region, September 2019 – May 2020. Points are individual mobility indicators derived from aggregated mobile device location data. Correlation coefficients are shown on the y-axis, and temporal lags (in weeks) between Rt and mobility are shown on the x-axis. Negative lags indicate behavior leads Rt, and positive lags indicate Rt leads behavior. The yellow shaded panel in each facet includes mobility indicators that have a leading, positive relationship with transmission, and hence would be considered predictive of transmission.

We used multivariable generalized additive models (GAMs) to measure non-linear relationships between mobility and Rt and model selection of GAMs to assess the relative importance of different indicators in predicting Rt during the 2019-2020 season, prior to the start of the COVID-19 pandemic (September 2019 – January 2020). For each pathogen, we allowed candidate models to include a smoothed temporal trend and up to two smoothed mobility terms. Across all pathogens, minimal models included a school-related behavioral indicator (foot traffic to schools or colleges) or the percentage of devices leaving home and a covariate related to large-scale population movement (between-neighborhood movement or out-of-state inflow), with the partial effects of most mobility covariates monotonically increasing with Rt (Figure S9).

### Initial effects of COVID-19 restrictions on mobility and respiratory virus circulation

The first SARS-CoV-2 infections in Washington state arose from a single introduction in late January or early February 2020, and at least one clade was circulating in the Seattle area for 3-6 weeks prior to February 28, when the first community acquired case was reported (*18*). To slow the spread of SARS-CoV-2, Washington declared a State of Emergency on February 29, closed schools in King, Pierce, and Snohomish counties on March 12, and enacted statewide stay-at-home (SAH) orders on March 23. In the interim, King County recommended that workplaces allow employees to work from home on March 4 and closed indoor dining and many other businesses on March 16.

Mobility levels declined substantially after February 29, and, by the start of King County’s business closures on March 16, foot traffic to transit stations were > 90% below baseline and out-of-state inflow and within-city mixing were > 60% below baseline (Figure 2, Figure S4). After the enactment of SAH orders on March 23, foot traffic to POIs and large-scale movements declined to 70-90% below baseline (Figure 2, Figure S4), while the percentage of devices staying completely at home increased to > 50% (Figure 2). Notably, social distancing measures altered not only the volume of movement between Seattle neighborhoods but also the presence and absence of connections between neighborhoods. Specifically, neighborhoods with low degree centrality (i.e., fewer than 10 connections to other neighborhoods) became much more prevalent compared to weeks prior to March 2020 (Figure 3).

The transmission rates of all respiratory pathogens dropped substantively after the State of Emergency, though some seasonal pathogens were already declining prior to February 29 (Figure 1, Figure S3). We measured the initial impacts of COVID-19 NPIs on respiratory virus circulation by comparing Rt values during the 2 weeks before and after the State of Emergency declaration on February 29 (Table 2). Early public health measures were effective at lowering SARS-CoV-2 transmission rates by 37% [95% CI: 25, 51]. Among endemic pathogens, influenza A/H1N1 virus, AdV, and hRV were the most impacted by pandemic-related behavioral changes, experiencing 29 – 42% reductions in transmissibility by March 16. hPIV 3 + 4, hMPV, and hCoV were also significantly affected, experiencing 20 – 25% declines in Rt. Reductions in RSV and influenza B virus were more modest, given their seasonal outbreaks had mostly concluded by late February. The hPIV 1 + 2 outbreak subsided in mid-February, and thus was not impacted by COVID-19 NPIs.

We observed strong relationships between mobility and the transmission of respiratory pathogens in the Spring of 2020. All mobility metrics were positive, leading indicators of Rt across all endemic viruses (Figure 4, Figure S10). In contrast, mobility lagged and was negatively correlated with SARS-CoV-2 transmission during this period (Figure 4, Figure S10). COVID-19 restrictions began to relax on May 4, 2020, when King County entered Phase 1 of the state’s reopening plan, allowing outdoor dining, worship services, and fitness centers to reopen with limited capacity. SARS-CoV-2 Rt values ranged from 0.8 to 0.9 throughout April and May and did not surpass 1 until late May (Figure 1). During late April and May, SARS-CoV-2 Rt became positively correlated and synchronous with most mobility indicators (r: 0.66 – 0.97) and inversely correlated with the stringency of NPIs (r: −0.97 – −0.81) (Figure 4, Figure S10). Yet, these relationships did not persist after the virus’s initial rebound in early June 2020, when King County reopened indoor dining and additional businesses (Figure S10). Due to the high degree of collinearity and concurvity among mobility metrics, we could not differentiate the effects of individual indicators on Rt during Seattle’s SAH period.

Population mobility did not immediately recover after SAH orders lifted in June 2020 (Figures 2-3, Figure S4). Visitor inflow from other WA counties and US states remained depressed at levels 50% below baseline until the spring and summer months of 2021, and within-and-between-neighborhood movement had not returned to pre-pandemic levels by the conclusion of our study in June 2022 (Figures 2-3, Figure S4). Further, SAH orders caused long-lasting structural changes to the mobility network of Seattle, wherein neighborhoods with high degree centrality (“hubs”) were most affected (Figure 3). After March 2020, neighborhoods with fewer than 10 connections to other neighborhoods became much more prevalent in the network, causing an overall downshift in the mobility network’s median degree for the remainder of the study period (Figure 3).

### Associations between SARS-CoV-2 transmission and behavior differ across COVID-19 waves

We measured cross-correlations between SARS-CoV-2 transmission, mobility, masking, and NPI stringency during subsequent pandemic waves in Seattle (Figure 5). After its first two COVID-19 outbreaks in Spring 2020 and Summer 2020, Seattle experienced a large third wave during winter 2020-2021 despite high masking rates (Figure 5). During November-December 2020, daily Rt was strongly correlated with foot traffic to restaurants (r: 0.65 – 0.83), the percentage of devices leaving home (r: 0.7 – 0.83), the influx of visitors from other WA counties and US states (r: 0.66 – 0.74), and NPI stringency (r: −0.85 – −0.76) (Figure 5). A smaller wave associated with the Alpha variant spanned March to May 2021, during which Rt was not significantly associated with mobility (Figure 5). COVID-19 cases and hospitalizations surged again in July 2021, due to the spread of the highly transmissible Delta variant. From late June to early August 2021, the percentage of residents not masking in public, the percentage of devices leaving home, and visitor inflow preceded or coincided with increases in Rt (r: 0.75 – 0.94) (Figure 5). Transmission decoupled from behavior during the Omicron BA.1 wave in late 2021, wherein SARS-CoV-2 Rt was negatively correlated and lagging most mobility metrics and positively correlated with masking and NPI stringency (the inverse of expected relationships) (Figure 5). Minimal GAMs fit to the first three months of each wave retained the percentage of devices leaving home, the percentage of residents not masking, foot traffic to restaurants, or out-of-state inflow (Figure S11), but associations between behavior and Rt were often negative.

**Figure 5.**
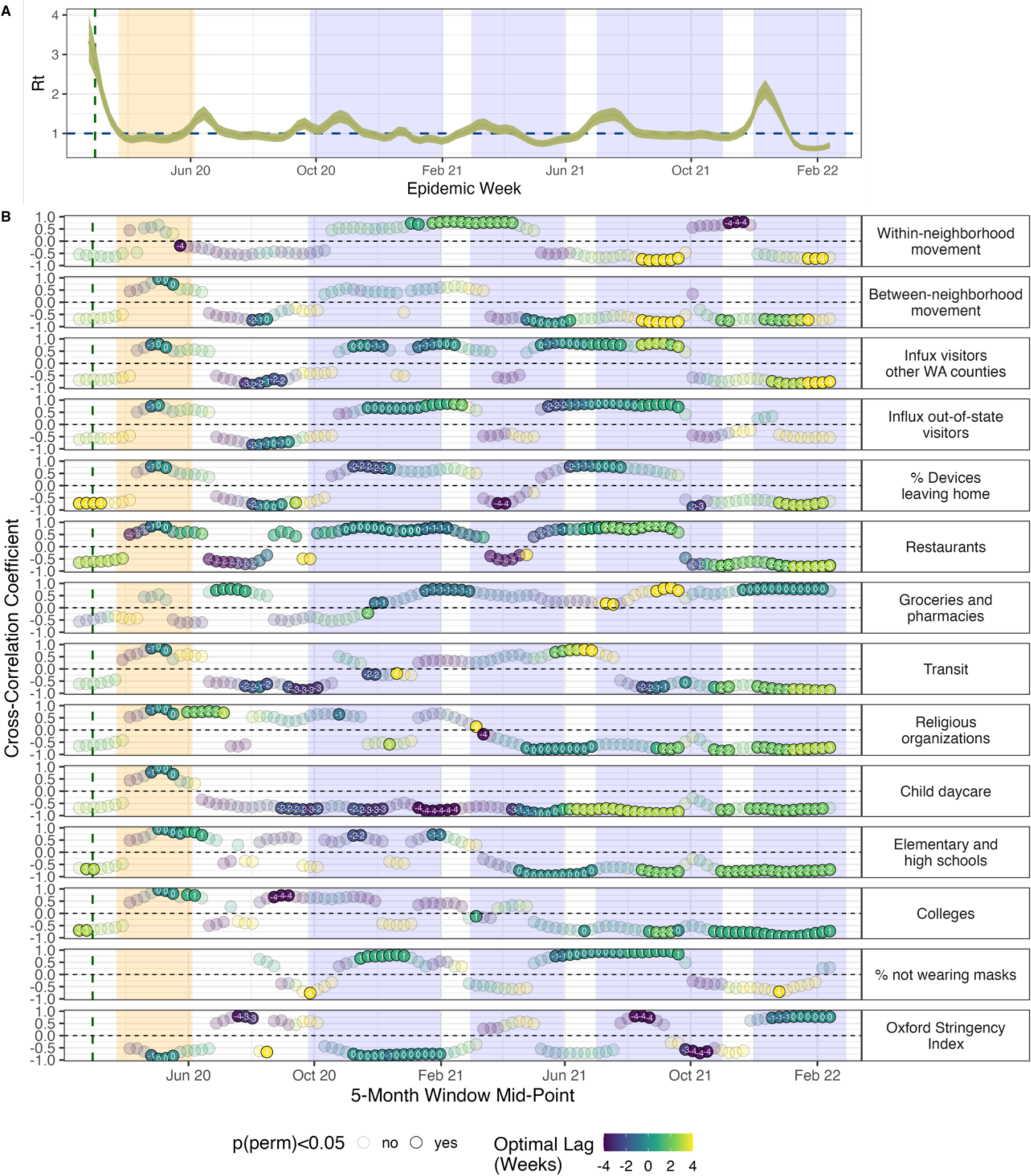
Relationships between mobility, masking, and SARS-CoV-2 transmission in the greater Seattle region during the COVID-19 pandemic, January 2020 – March 2022. **A.** Weekly effective reproduction number (Rt) of SARS-CoV-2, and **B.** Rolling cross-correlations between weekly Rt and mobility and behavioral indicators. Points represent the maximum coefficient values for 5-month rolling cross-correlations between weekly Rt and individual mobility metrics. Point color and the number within each point indicate the lag in weeks corresponding to the maximum cross-correlation coefficient value for each 5-month period (“optimal lag”). Negative values indicate that behavior leads Rt, and positive values indicate that Rt leads behavior. A lag of 0 indicates that the time series are in phase (i.e., synchronous). Point transparency indicates statistical significance of the cross-correlation coefficient (yes: solid, no: transparent). The vertical green dashed line indicates the date of Washington’s State of Emergency declaration (February 29, 2020), and the vertical orange shaded panel indicates Seattle’s stay-at-home period (March 23 – June 5, 2020). The blue shaded panels indicate the timing of four COVID-19 waves in Seattle: the winter 2020-2021 wave, the Alpha wave in Spring 2021, the Delta wave in Summer 2021, and the Omicron BA.1 wave during late 2021 to early 2022.

### Mobility predictors of endemic pathogen rebound during the COVID-19 pandemic

We observed a remarkably fast rebound of hRV, and AdV to a lesser degree, when SAH restrictions relaxed in early June 2020 (Figure 1, Figures S2-S3). Increases in hRV transmission were preceded by or synchronous with the percentage of devices leaving home, foot traffic to restaurants, between-neighborhood movement, and visitor inflow from June to early August 2020 (r: 0.75 – 0.96), inversely correlated with NPI stringency during September 2020 (r: −0.79 – −0.69), and continuously correlated with foot traffic to religious organizations until late 2020 (r: 0.64 – 0.91) (Figure S12). The rebound of AdV was preceded by or synchronous with slight increases in foot traffic to schools and religious organizations from June to early August 2020 (r: 0.7 – 0.87) (Figure S12). For both viruses, minimal GAMs fit to the first six months of rebound retained the percentage of devices leaving home and out-of-state inflow (Figure S13).

Resurgence of other endemic viruses, including hCoV, hPIV, hMPV, and RSV B, was not observed until early-to-mid 2021, and epidemic peaks for hCoV, hMPV, and RSV B occurred during the spring or summer outside of their typical seasons (Figure 1, Figures S2-S3). We measured univariate associations between mobility, masking, NPI stringency, and daily transmissibility and found fewer clear relationships compared to Seattle’s SAH period and the 2019-2020 winter season. Multiple mobility indicators preceded or coincided with the rebound of these viruses, though, for RSV B and hCoV, associations were briefer compared to those observed during seasonal outbreaks prior to the pandemic (Figure S14). For example, at the beginning of RSV B’s rebound in summer 2021, increases in Rt were preceded by between-neighborhood movement and foot traffic to schools, child daycares, and religious organizations (r: 0.7 – 0.9) and synchronous with the percentage of devices leaving home, the percentage of residents not masking in public, and out-of-state inflow (r: 0.7 – 0.91) (Figure S14). These associations were more transient compared to the pre-pandemic period, persisting for 1-2 months instead of 2-3 months (Figures S8 and S14). Minimal GAMs fit to the first three months of each virus’s rebound retained between-neighborhood movement and out-of-state inflow, though relationships between mobility and Rt were nonlinear and not consistently positive (Figure S15).

In late 2021, endemic virus circulation declined as Omicron cases surged, and reductions in endemic virus transmission were universally preceded by or coincided with drops in mobility (Figure S12, Figure S14, Figure S16). For example, reductions in most mobility indicators preceded declines in RSV B, hMPV, hCoV, and hPIV 3 + 4 circulation by 1 to 4 weeks, while reduced visits to child daycares, schools, colleges, and transit stations were synchronous with declines in hRV transmission (Figures S12, Figure S14). The transmission rates of RSV B, hMPV, and hCoV were also associated with the percentage of devices staying home, which spiked from 35% to 50% in December 2021 (Figure 2, Figure S16). During the Omicron wave (November 2021 – January 2022), the best fit minimal GAMs for each virus retained a school-related behavioral indicator, the percentage of devices leaving home, or out-of-state inflow, similar to results observed for the 2019-2020 season (Figure S17).

Although our study’s aim was inferential rather than predictive, we built forecasting models of daily Rt for three viruses that circulated continuously throughout the study period: hRV, AdV, and SARS-CoV-2. We used mobility metrics, the co-circulation of other viruses, and activity of the target virus during the previous two weeks (14 autoregressive terms) as candidate predictors. For each virus, models with only mobility terms did not outperform models with only autoregressive terms (Figures S18-S20) but still produced accurate forecasts over the entire study period (Pearson’s r with observed data, r > 0.8), and especially during Seattle’s stay-at-home orders and the initial lifting of restrictions (r: 0.94 – 0.95) (Table S3). Methodological details and results are provided in the supplementary material.

## Discussion

We investigated the impacts of human behavior on the transmission of respiratory viruses in the greater Seattle region, during pre- and post-pandemic years, by modeling incidence derived from hospital and community-based respiratory surveillance and human movements from high-resolution mobile device location data. From November 2018 to June 2022, we characterized the epidemiological dynamics of 16 endemic viruses and SARS-CoV-2 and related changes in daily transmissibility (time-varying effective reproduction numbers, Rt) to trends in population mobility, masking, and COVID-19 non-pharmaceutical interventions (NPIs). To our knowledge, this is the first study to explore the effects of mobility and behavior on transmission across a large set of endemic pathogens; interestingly, we saw notable heterogeneity in the timing and size of each endemic pathogen’s rebound during the pandemic period.

Mobility was most predictive of transmission during periods of dramatic behavioral change, as observed during Seattle’s stay-at-home (SAH) orders in March 2020. Smaller-scale changes in mobility also preceded or coincided with increases in Rt at the beginning of outbreaks and declines in Rt during shorter interruptions to human movement, as observed during a major snowstorm in February 2019 and the Omicron BA.1 wave in late 2021. Across all endemic viruses, trends in daily Rt were repeatedly associated with the same set of mobility metrics, including foot traffic to elementary and high schools, colleges, child daycare centers, restaurants, and religious organizations, the percentage of devices leaving home, between-neighborhood movement, and the inflow of visitors from other WA counties and US states. Outside the SAH period, SARS-CoV-2 transmission correlated with foot traffic to restaurants and colleges, the percentage of devices leaving home, and visitor inflow. Foot traffic to specific businesses and educational and religious activities may approximate close contacts or crowded conditions that facilitate direct, aerosol, or droplet transmission, while the percentage of devices leaving home, within-city mixing, and inflow from other regions may be indicative of human movements that promote viral introductions and dispersal.

The age distribution of infections may explain slight differences in which categories of POIs correlated with endemic virus transmission versus SARS-CoV-2 transmission. Recurrent associations between endemic virus Rt and visits to schools and daycares are consistent with children experiencing the highest rates of (endemic) respiratory infections and schools and daycares acting as a major source of transmission in the community (*29–33*). Unlike endemic respiratory viruses, all age groups are susceptible to SARS-CoV-2 infection. Correlations between SARS-CoV-2 Rt and foot traffic to colleges and restaurants, but not schools or daycares, could be attributed to greater rates of symptomatic infection (and hence shedding propensity) in adults relative to younger age groups (*34*) or to the greater relevance of adult networks in spreading SARS-CoV-2 compared to endemic viruses.

COVID-19 NPIs significantly perturbed the transmission of respiratory pathogens at a global level (*1–8*), causing the complete disappearance, delayed return, or “off-season” outbreaks of endemic pathogens (*6–9*). In Seattle, all endemic respiratory viruses experienced rapid declines at the beginning of Seattle’s SAH orders in March 2020, but, as restrictions eased, their rebound was heterogeneous. Similar to trends observed in the US and other countries (*4, 6, 7, 10, 35–39*), the circulation of hRV and AdV resumed in early June 2020, immediately after nonessential businesses and indoor dining reopened, while other respiratory viruses virtually disappeared during March 2020 and did not recirculate until 2021. Further, the resurgence of RSV B, hCoV, and hMPV occurred outside of their typical seasons, as reported in other locations (*7–9*). After the initial easing of COVID-19 restrictions, relationships between endemic virus dynamics and mobility were less clear compared to Seattle’s SAH orders or the 2019-2020 winter season, potentially due to continued social distancing and masking, a more refined understanding of “high risk” activities, the delay of in-person instruction for school students until spring 2021, or structural changes to Seattle’s mobility network. Nonetheless, associations between endemic virus Rt and population behavior were overall stronger and longer-lasting than those observed for SARS-CoV-2.

It is remarkable that the two viruses that rebounded immediately after lockdown restrictions lifted, hRV and AdV, are non-enveloped viruses, while the other viruses studied here are enveloped. The immediate rebound of non-enveloped viruses could be attributed to viral stability and persistence. Non-enveloped viruses are less susceptible to lipophilic disinfection and can persist on hands and fomites for longer periods of time than enveloped viruses (*40, 41*). In addition of the presence or absence of an envelope, several other factors, such as transmission mode, seasonality, source/sink dynamics, and duration of infectious period and immunity, could have affected the timing of rebound. While enveloped viruses disappeared in March 2020, non-enveloped viruses may have continued to spread during SAH restrictions, due to their longer periods of viral shedding, high preexisting community prevalence, or ability to persist on environmental surfaces (*36, 40–42*). We hypothesize that low levels of transmission or residual viral particles on surfaces, combined with slight increases in movement, close contacts, and visitor inflow, were sufficient to facilitate the rapid rebound and ongoing transmission of hRV and AdV after SAH orders lifted in June 2020. Further, surgical masks are less effective at filtering hRV compared to influenza viruses and seasonal coronaviruses (*43*).

Our study period encompasses the two respiratory virus seasons prior to the start of the COVID-19 pandemic and two pandemic years. The Seattle Flu Study (SFS) began collecting samples in November 2018, which precluded us from evaluating potential leading indicators of transmission at the beginning of the 2018-2019 season. However, we were able to detect strong links between mobility and transmission in February 2019 when a major snowstorm forced work and school closures, consistent with previous SFS research that did not specifically examine cell phone mobility patterns (*21*). SFS continued to collect respiratory samples throughout 2019, enabling us to test for leading indicators of transmission during the 2019-2020 winter season. During Fall 2019, the transmission dynamics of enveloped viruses were more strongly correlated with mobility than those of non-enveloped viruses. For enveloped viruses, foot traffic to schools and colleges, between-neighborhood movement, and visitor inflow preceded or coincided with increases in transmission, with associations between Rt and mobility weakening over the course of the season, presumably due to accumulating immunity in the population. During this same period, non-enveloped viruses had fewer positive relationships with mobility, potentially because hRV and AdV circulate year-round and have less defined peaks and troughs.

SARS-CoV-2 began circulating in the greater Seattle region during January or February 2020 (*18*), with the first community-acquired case confirmed on February 28, 2020. Mobility had a negative, lagging relationship with SARS-CoV-2 Rt during the early months of 2020, suggesting that Seattle residents adjusted their behavior in response to COVID-19 case counts or restrictions rather than the reverse. Mobility was briefly predictive of SARS-CoV-2 transmission when social distancing restrictions first relaxed in summer 2020 and during the winter 2020-2021 and Delta waves, before ultimately decoupling from Rt during the Omicron BA.1 wave in late 2021. Compared to prior variants of concern (VOCs), Omicron BA.1 has a shorter serial interval, increased immune evasion, and greater intrinsic transmissibility (*44–46*). A phylogeographic study linked inter-city travel to the spatial spread of individual Omicron BA.1 lineages in the UK (*47*), suggesting that daily Rt estimated from all COVID-19 cases combined may lack the resolution to relate the rapid spread of Omicron BA.1 to mobility patterns (*see methods for comments on VOC-specific Rt analyses*).

Climate affects the stability and seasonal dynamics of respiratory viruses (*48–51*) but was an unlikely driver of endemic virus resurgence in Seattle. hRV and AdV have year-round circulation with peaks in the spring and early autumn (*52, 53*), while influenza viruses, RSV, hCoV, hMPV, and hPIV have distinct seasonality with peaks during the winter or spring (*53–56*). The lifting of SAH orders in June 2020 coincided with the typical timing of low circulation for enveloped viruses and increasing activity for non-enveloped viruses. However, the intersection of relaxing NPIs with warmer weather cannot account for the global differences observed between non-enveloped and enveloped virus rebound. The prolonged absence of influenza and RSV circulation was also reported during the Southern Hemisphere winter (*2, 5, 9*), and climatic factors cannot explain the rebound of hCoV, hMPV, and RSV B outside of their typical seasons.

Our findings suggest that in-person school instruction played a key role in the rebound of enveloped viruses in Seattle. Prior research has shown that increased contact rates among older children during school terms influence the timing of seasonal influenza and “common cold” virus outbreaks (*29, 57, 58*), and younger children and adults acquire influenza and RSV infections from preschool or school-aged children living in the same household (*59–61*). All King County public school districts began the 2020-2021 academic year remotely (*62*), with some districts offering limited in-person instruction to special education students during Fall 2020. Foot traffic to schools was 75% below baseline in Fall 2020, gradually increased to 50% below baseline during Spring 2021, and returned to baseline levels in Fall 2021. We observed that the circulation of hCoV, hPIV, and hMPV increased after elementary school students were offered in-person instruction in February and March 2021 and that the off-season RSV B wave in summer 2021 began directly after hybrid learning became available to all grades in mid-April (*62*). These trends suggest that a year of remote learning, and in turn reduced contacts among school-aged children, contributed to the delayed return of enveloped virus circulation to Seattle.

The rebound of enveloped viruses also coincided with increasing rates of travel into Seattle. Annual influenza epidemics in North America are seeded via air travel by strains originating in East and Southeast Asia (*63, 64*), and the regional spread of influenza viruses correlates closely with work commutes (*65, 66*). We did not have data on international air travel or commuting patterns, but cell phone data show that the inflow of visitors from other WA counties and US states was 50% below baseline throughout 2020 and did not return to pre-pandemic levels until late spring or summer 2021. Although the contribution of local persistence versus external seeding is less understood for other seasonal respiratory viruses, increasing inflow into Seattle likely imported cases from other regions, seeding new outbreaks (*9*).

Lastly, prolonged lack of exposure due to reduced viral circulation during 2020 and 2021 is expected to have increased the cohort of children completely naïve to various respiratory viruses and the waning of immunity in previously infected individuals (*14, 67*). This “immunity debt” may have provided enough susceptible individuals to sustain spring and summer outbreaks of enveloped viruses. Although we expected these outbreaks to be larger or more severe than those observed during pre-pandemic seasons, substantial influenza and RSV epidemics did not occur until the Fall of 2022, potentially due to Seattle residents continuing to social distance and mask throughout 2021 or negative interference between Omicron BA.1 viruses and endemic viruses during the 2021-2022 winter season. After the conclusion of our study, the 2022-2023 season saw atypically early outbreaks of influenza and RSV and higher hospitalization rates in children and adolescents compared to pre-pandemic seasons (*15, 68*).

Our study has limitations related to the type of cell phone mobility data used and the underlying demographics of mobile device data in general. Young children experience the highest rates of endemic respiratory infections, but SafeGraph does not track individuals younger than 16 years of age. Nonetheless, we found that visits to schools and daycares were positive, leading indicators of transmission, both prior to and during the pandemic, demonstrating that mobile phone data derived from adults can approximate the movements or contacts of children. Second, although we found statistically significant associations between foot traffic indicators and pathogen transmission, spatial co-location data may better approximate the interpersonal contacts that underlie transmission and reduce statistical noise. Longitudinal cross-sectional surveys on social interactions, such as the CoMix survey in England, can provide more direct measures of epidemiologically relevant behavior and more representative samples of populations than mobile device data (*69*). However, to our knowledge, similar data do not exist for the US.

Our findings are subject to several other limitations. First, due to the limited number of seasons in our study, we could not determine if leading indicators of transmission are consistent across timespans longer than four years. Although SFS continued to conduct respiratory surveillance into the 2022-2023 winter season, its community surveillance approach changed substantially after July 2022, making it difficult to extend our study. Second, variability in test volume over time caused SFS surveillance to sometimes miss less prevalent pathogens. For example, SFS detected only a few cases during a small influenza A/H3N2 wave in winter 2021-2022. Third, our multiplex PCR assay could not distinguish between types, strains, or serotypes of some pathogens (hCoV, hPIV, AdV, hRV). Consequently, our Rt estimates may average over heterogeneities in transmission dynamics among viruses belonging to the same species (*19*). Fourth, previous work has shown that SARS-CoV-2 transmission dynamics differed between North and South King County (*20, 70*), potentially due to socioeconomic inequities (e.g., differences in income, household size, proportion of essential workers) and North King County maintaining a greater reduction in mobility over time. However, we did not have sufficient surveillance data to explore geographic differences in the transmission dynamics of endemic viruses. Fifth, population immunity may modulate relationships between mobility and transmission; analysis of serologic data could shed light on this question. Additional research is needed to delineate the contributions of an increasingly susceptible population and decreased social distancing to the rebound of endemic viruses.

In summary, mobility patterns are most predictive of respiratory virus transmission during drastic changes in contacts, and, to a lesser extent, at the beginning of epidemic waves. During the pandemic period, endemic respiratory viruses exhibited stronger relationships with mobility than pandemic SARS-CoV-2. As SARS-CoV-2 transitions to endemicity, relationships with mobility could gradually start to operate similarly to those of other enveloped viruses. Our study shows that mobile phone data can approximate transmission relevant contacts and has the potential to support the surveillance of endemic respiratory viruses, with the caveat that relationships between transmission and mobility vary depending on the pathogen, magnitude of mobility change, and phase of the epidemic. Future research should consider other host factors, such as prior immunity, and more direct proxies of interpersonal contacts to further disentangle relationships between population behavior and respiratory virus dynamics.

## Methods

### Virologic surveillance and laboratory methods

This population-level study uses cross-sectional surveillance data collected through the Seattle Flu Study (SFS) from November 2018 to June 2022. Initiated in November 2018, SFS was a multi-arm surveillance study of influenza and other respiratory pathogens in the greater Seattle, WA region, that utilized community and hospital-based sampling (*17*). In its first 1.5 years, SFS tracked the transmission of influenza and other respiratory pathogens in the Seattle region by testing swabs collected at hospitals, community sites (e.g., kiosks in high foot traffic areas, outpatient clinics, workplaces, college campuses), and through its swab-and-send at-home testing study (*17, 22*) (Table S2). The SFS team launched the greater Seattle Coronavirus Assessment Network (SCAN) in March 2020 to detect and understand the spread of SARS-CoV-2 (*23*). SCAN deployed self-administered at-home testing kits to monitor the spread of both SARS-CoV-2 and endemic respiratory pathogens from March 2020 to July 2022. We describe each surveillance arm in the supplementary methods.

The Institutional Review Board of the University of Washington gave ethical approval of this work (protocol #00006181). All participants who contributed specimens to the Seattle Flu Study or Greater Seattle Coronavirus Assessment Network provided informed consent at enrollment. Informed consent for residual sample and clinical data collection was waived, as these samples were already collected as part of routine clinical care, and it was not possible to re-contact these individuals. This study followed the Strengthening the Reporting of Observational Studies in Epidemiology (STROBE) reporting guidelines for cross-sectional studies.

Each respiratory specimen was screened in duplicate for a panel of respiratory pathogens using a custom TaqMan RT-PCR OpenArray panel (Thermo Fisher). Laboratory methods are described in detail elsewhere (*20, 22*). Pathogen targets included adenovirus (AdV); human bocavirus (hBoV); human coronaviruses (hCoV) 229E, OC43, HKU1, and NL63; human metapneumovirus (hMPV); human parainfluenza viruses (hPIV) 1, 2, 3, and 4; human parechovirus (hPeV); influenza A (IAV) H1N1 and H3N2; pan influenza A (IAV); pan influenza B (IBV); pan influenza C (ICV); respiratory syncytial viruses (RSV) A and B; human rhinovirus (hRV); enterovirus D68 (EV.D68); pan enterovirus excluding D68 (EV); *Streptococcus pneumoniae* (Spn); *Mycoplasma pneumoniae* (Mpn); *Chlamydia pneumoniae* (Cpn); and SARS-CoV-2. Due to assay limitations, epidemiologically distinct strains were grouped into one assay each for hCoV 229E and hCoV OC43, hCoV HKU1 and hCoV NL63, hPIV 1 and hPIV 2, hPIV 3 and hPIV 4, EV, hRV, and AdV.

A substantial number of specimens tested positive for SPn, a common commensal, with SPn detected in 27.3% of positive samples prior to March 2020 and 18.7% of positive samples after March 2020 (Figure S2). We opted to not include SPn in downstream analyses due to our inability to distinguish acute infections from chronic carriage.

### Syndromic surveillance data

We obtained respiratory syndromic surveillance data for King County, WA from the Rapid Health Information Network (RHINO) program at the Washington Department of Health (WA DOH) (Figure S21). Syndrome criteria are defined by the Electronic Surveillance System for the Early Notification of Community-Based Epidemics (ESSENCE). We received weekly counts of total emergency department (ED) visits and ED visits classified as *influenza-like illness* (ILI) (mention OR diagnosis of influenza OR fever (≥100°F) and cough OR fever (≥100F) and sore throat), *COVID-like illness* (CLI) (mention OR diagnosis of coronavirus AND no diagnosis of influenza OR fever OR chills AND cough OR shortness of breath OR difficulty breathing), and *broad respiratory illness* (acute bronchitis OR chest congestion OR cough OR difficulty breathing OR hemoptysis OR laryngitis OR lower respiratory infection OR nasal congestion OR otitis media OR pneumonia OR shortness of breath OR sore throat OR upper respiratory infection OR wheezing OR acute respiratory distress). Weekly data were disaggregated by age group (0-4, 5-24, 25-64, and ≥65). We collapsed age groups into two categories, < 5 and ≥ 5 years of age, and calculated the weekly proportion of ED visits coded as ILI, CLI, or broad respiratory illness (Figure S21).

Respiratory syndromic surveillance data for Washington state were obtained from the U.S. Outpatient Influenza-like Illness Surveillance Network (ILINet) via the CDC FluView Interactive dashboard (*71*). ILINet consists of approximately 3,200 sentinel outpatient healthcare providers throughout the United States that report the total number of consultations for any reason and the number of consultations for ILI every week. ILI is defined as fever (temperature of 100°F [37.8°C] or greater) and a cough and/or a sore throat. ILINet provides the weekly proportion of outpatient consultations for ILI and the number of ILI encounters by age group (0-4, 5-24, 25-64, and ≥65).

### Data on cell phone mobility, masking, and the stringency of non-pharmaceutical interventions

We obtained mobile device location data from SafeGraph (https://safegraph.com/), a data company that aggregates anonymized location data from 40 million devices, or approximately 10% of the United States population, to measure foot traffic to over 6 million physical places (points of interest) in the US. We estimated foot traffic to specific points of interests (POIs), movement within and between census block groups, and the in-flow of visitors residing outside of King County from June 2018 to June 2022, using SafeGraph’s “Weekly Patterns” dataset, which provides weekly counts of the total number of unique devices visiting a POI from a particular home location. POIs are fixed locations, such as businesses or attractions. A “visit” indicates that a device entered the building or spatial perimeter designated as a POI. A “home location” of a device is defined as its common nighttime (18:00-7:00) census block group (CBG) for the past 6 consecutive weeks. We restricted our datasets to King County POIs that had been recorded in SafeGraph’s dataset since January 2019.

To measure movement within and between CBGs (“neighborhoods”) in King County, we extracted the home CBG of devices visiting points of interest (POIs) and limited the dataset to devices with home locations in the CBG of a given POI (within-neighborhood movement) or with home locations in CBGs outside of a given POI’s CBG (between-neighborhood movement). To measure the inflow of visitors from other counties in Washington state or from out-of-state, we limited the dataset to devices visiting POIs in King County with home locations in other WA counties or in other US states, respectively. For each POI in each week, we excluded home census block groups with fewer than five visitors to that POI. To measure foot traffic to specific categories of POIs, we aggregated daily visits to POIs by North American Industry Classification System (NAICS) category, without considering the home locations of devices visiting these POIs. To adjust for variation in SafeGraph’s device panel size over time, we divided Washington’s census population size by the number of devices in SafeGraph’s panel with home locations in Washington state each month and multiplied the number of daily or weekly visitors by that value. For each mobility indicator, we summed adjusted daily or weekly visits across POIs and measured the percent change in movement over time relative to the average movement observed in all of 2019, excluding national holidays.

Daily data on the percentage of devices staying home in King County were obtained from SafeGraph’s Social Distancing Metrics (https://docs.safegraph.com/docs/social-distancing-metrics) and Meta Data for Good’s Movement Range Maps (https://dataforgood.facebook.com/dfg/tools/movement-range-maps) (*72*). SafeGraph social distancing metrics were available from January 1, 2019, to April 16, 2021, and Meta Movement Range Maps were available from March 1, 2020, to May 22, 2022. Trends in the percentage of devices staying home were almost identical across the two data sources, though the percentage of devices staying home in the Meta dataset was lower than that observed in the SafeGraph dataset. We added a scaling factor to the Meta indicator and joined the two time series to create a single metric for our study period.

We obtained survey data on the daily percentage of King County residents wearing face masks in public from the Carnegie Mellon University Delphi Group Covidcast API (*28*). Masking data were collected as part of the COVID-19 Trends and Impact Survey conducted by the Delphi group, in collaboration with Meta and a consortium of universities and public health officials (*73*). The survey ran continuously from April 6, 2020, to June 25, 2022, with approximately 40,000 individuals in the United States participating every day. The survey included specific questions about masking from September 8, 2020, to June 25, 2022. We supplemented the Covidcast King County masking data with COVIDNearYou survey data for Washington state (*74*) to extend the time series to June 2, 2020.

We extracted data collected by the Oxford COVID-19 Government Response Tracker (OxCGRT)(*27*) to measure variation in Washington state’s government policies related to COVID-19 from March 1, 2020 to June 30, 2022. The OxCGRT database tracked publicly available information for policies related to closure and containment, health, and economic policy in 180 countries, recording policy responses on ordinal or continuous scales for 19 policy areas. We obtained daily values for the stringency index, which combines all containment and closure (C) indicators and the H1 indicator (public information campaigns).

### Reconstructing pathogen incidences

While SFS sampling is robust enough to provide granular (daily) surveillance data on the circulation of multiple pathogens, the diversity of SFS sampling schemes requires pre-processing to infer pathogen incidence. To properly reconstruct pathogen incidences through time, we considered the different populations sampled by SFS, particularly regarding age group, clinical setting, and the presence of respiratory symptoms.

We first excluded samples with missing age or home address information (as reported by individuals participating in community surveillance or obtained through electronic hospital records), samples from individuals residing outside the greater Seattle region (King, Pierce, Snohomish, Kitsap, San Juan, Whatcom, Skagit, Island, Clallam, Jefferson, Mason, and Thurston counties), samples from individuals who were asymptomatic for respiratory illness, and samples from multiple testing of individuals. If an individual was tested more than once in a 30-day period, we kept one result per pathogen in that period. If test results for all pathogens were consistent across the testing instances in the 30-day period, we kept the results from the first testing instance and discarded the subsequent instances. If an individual tested negative and then positive, or tested positive then negative, we kept the result for the first positive testing instance and discarded the instances prior to or after that result. We also excluded samples collected as part of Public Health – Seattle & King County’s (PHSKC) contact tracing efforts or through collaborations with community-based organizations.

Next, we used a three-step approach to control for sampling variation over time. In the first step, we disaggregated daily pathogen presence and absence data derived from OpenArray testing by clinical setting (hospital or community) and age (≥ 5 years or < 5 years). We then divided the number of positive samples for each pathogen by the total number of specimens tested in each setting and age stratum. Daily proportion positive values were multiplied by the expected age distribution of cases for each pathogen in each setting, which were obtained from the U.S. Outpatient Influenza-like Illness Surveillance Network (ILINet) (*71*), the U.S. Influenza Hospitalization Surveillance Network (FluSurv-NET)(*75*), the Washington State Department of Health (*76*), or published literature (Table S4).

In the second step, we combined pathogen proportion positive information from SFS virologic surveillance with citywide syndromic surveillance indicators for respiratory illnesses. A similar approach has been successfully used to model influenza activity over multiple seasons (*77*). Specifically, we multiplied the age-adjusted proportion positive data by a weekly indicator of the proportion of the King County population seeking care for respiratory illness at emergency departments (ED). This adjustment was applied separately to community and hospital data to generate community and hospital-based incidences for each pathogen. For hospital- and community-based incidences of each pathogen, we multiplied daily proportion positive values for individuals < 5 or ≥ 5 years by the weekly percentage of ED visits coded as general respiratory illness (all endemic viruses except influenza), influenza-like illness, ILI (influenza), or COVID-like illness, CLI (SARS-CoV-2) for each age group.

In a third step, hospital- and community-based incidences were each rescaled to fall between 0 and 1 and summed to provide an aggregate measure of incidences in the greater Seattle region. We used this approach to estimate daily incidences from November 2018 to June 2022 for pathogens with sufficient sampling (≥ 400 positive specimens during 2018 – 2022), including influenza A/H1N1, A/H3N2, and B viruses, RSV A and B, seasonal coronaviruses, human parainfluenza viruses, human metapneumovirus, rhinovirus, and adenovirus.

We estimated daily SARS-CoV-2 incidence from publicly available COVID-19 case data for King County (*76*) because SCAN did not test respiratory specimens for SARS-CoV-2 during May and June 2020 (Figure S22).

### Statistical analysis

All analyses were performed using R version 4.3.0.

#### Transmission modeling

For each pathogen, we estimated time-varying (instantaneous) reproduction numbers, Rt, by date of infection using the Epidemia R package (*26*). The instantaneous reproduction number is the number of secondary cases arising from a symptomatic individual at a particular time, assuming conditions remain identical after that time. Epidemia implements a semi-mechanistic Bayesian model using the probabilistic programming language Stan (*78*). Prior to Rt estimation, we computed proxies of daily case counts of endemic pathogens by multiplying daily incidence rates by 1000 and rounding the resultant values to integers. We estimated reporting delays (i.e., the delay from symptom onset to testing) using kiosk, swab-and-send, and SCAN questionnaire metadata collected from symptomatic individuals who tested positive for endemic respiratory viruses (hRV, N = 4848 survey responses; influenza viruses, N = 830; RSV, N = 423; hPIV, N = 325; hCoV, N = 666; hMPV, N = 148; AdV, N = 443) or SARS-CoV-2 (N = 3566). We used Stan to fit a lognormal distribution to 100 subsampled bootstraps (each with 250 samples drawn with replacement) of the available reporting delay data separately for all endemic respiratory viruses combined and SARS-CoV-2, with a maximum allowed delay of 30 days (EpiNow2 R package (*79*)). This resulted in a lognormal onset-to-testing delay distribution with mean 0.49 (1.02 SD) days for endemic viruses and mean 0.65 (1.1 SD) days for SARS-CoV-2.

To estimate Rt, observed cases were modelled as a function of latent infections in the population, assuming a negative binomial distribution. For each pathogen, we estimated the time distribution for infection-to-case-observation by summing the lognormal-distributed incubation period and the lognormal-distributed reporting delay. Pathogen-specific incubation periods and generation or serial intervals were obtained from published literature (Table S5). Instead of using the renewal equation to propagate infections, we treated infections as latent parameters in the model, because the additional variance around infections leads to a posterior distribution that is easier to sample (*26*). To control for temporal autocorrelation, we modelled Rt as a daily random walk. Epidemic trajectories were fit independently using Stan’s Hamiltonian Monte Carlo sampler. For each model, we ran 4 chains, each for 30,000 iterations (including a burn-in period of 15,000 iterations that was discarded), producing a total posterior sample size of 60,000. We verified convergence by confirming that all parameters had sufficiently low ^R^^ hat values (all R hat < 1.1) and sufficiently large effective sample sizes (>15% of the total sample size).

Following methods from (*1*), we evaluated changes in transmissibility during the two weeks before and after two major events during our study period: a major snowstorm in February 2019 and the initiation of COVID-19 social distancing measures in March 2020. We used t-tests for the ratio of two means to compare Rt values before and after each event and Fieller’s theorem to calculate the 95% confidence intervals of changes in Rt.

#### Cross-correlations between human behavior and pathogen transmission

From Fall 2019 to Summer 2022, we computed Pearson cross-correlations between weekly pathogen specific Rt values and the weekly percent change from baseline in mobility, in rolling five-month windows. During the 2018-2019 respiratory virus season, we computed cross-correlations between daily Rt values and the daily percent change from baseline in mobility in rolling one-month windows, due to limited data at the start of that season (SFS began collecting samples in November 2018) and to better capture the effects of the 12-day snowstorm in February 2019. For time periods that data were available, we estimated weekly cross-correlations and optimal lags between Rt and the proportion of individuals masking in public (June 2020 to June 2022) and between Rt and the Oxford Stringency Index (March 2020 to June 2022). In all analyses, cross-correlations were weighted with an exponential decay such that observations at the edges of each time window were weighted approximately 50% less than observations at the window midpoint.

For each rolling window, we estimated weighted cross-correlations between mobility behavior and Rt at different lags (up to 4 weeks for 5-month rolling windows and up to 14 days for one-month rolling windows) and extracted the maximum (absolute) coefficient value and the lag (in weeks or days) at which this value occurred (the ‘optimal lag’). Negative lag values indicate behavior leads Rt, and positive lag values indicate Rt leads behavior. A lag of 0 indicates that two time series are in phase (i.e., synchronous). To generate monthly cross-correlations and lags, we averaged the correlation coefficients and optimal lags of window midpoints that fell within a given calendar month. As an example, for five-month rolling windows each month’s statistics are an average of correlation coefficients and lags for dates falling 10 weeks prior to and 10 weeks following each week in that month. To test the statistical significance of cross-correlations for each rolling window, we used a block bootstrap approach to generate 1000 samples of each mobility time series in two week increments and recomputed weighted cross-correlations between Rt and mobility for each replicate, yielding a null distribution of 1000 cross-correlations. Cross-correlations between Rt and mobility indicators were considered statistically significant when observed coefficients were outside the bounds of the null distribution’s 90% interval.

As a sensitivity analysis, we estimated the daily transmissibility of the ancestral SARS-CoV-2 virus and each major variant of concern (VOC), using generation intervals, incubation periods, and reporting delays specific to each lineage, and computed rolling cross-correlations between VOC-specific Rt values and behavioral indicators. Most VOC time series were too short to measure dynamic changes in correlations between Rt and behavior, likely because we could not include the period immediately preceding increases in Rt in VOC-specific analyses.

#### Multivariable generalized additive regression models

We used multivariable generalized additive regression models (GAMs) to measure non-linear relationships between mobility and Rt and assess the relative importance of different indicators in predicting Rt for each pathogen during key epidemiological timepoints: the beginning of the 2019-2020 respiratory virus season (September 2019 – January 2020), the first three months of each of four COVID-19 waves, the first six months of rebound of non-enveloped viruses (June – November 2020), the first three months of rebound of each enveloped virus in 2021 (January – August 2021), and the decline of endemic viruses during the Omicron wave in late 2021 (November 2021 – January 2022). We used the mgcv R package (*80*) to fit each GAM with a Gamma error distribution and log link. Mobility covariates and time trends were modelled using thin plate regression splines (the default smooth for s terms). We specified for the model to add an extra penalty to each term so that it could be penalized to zero (select = TRUE), which enables the smoothing parameter estimation to completely remove terms when fitting the model.

To further refine our set of predictors and reduce concurvity, we used Akaike’s Information Criteria corrected for small sample sizes (AICc) to select the best fit “minimal” model for each pathogen, allowing candidate models to include a smoothed weekly time trend and up to two smoothed population behavior terms. Candidate predictors included within-neighborhood movement, between-neighborhood movement, inflow from other WA counties, inflow from other US states, the percentage of devices leaving home, and foot traffic to different categories of POIs, including restaurants, religious organizations, child daycare centers, elementary and high schools, and colleges. For SARS-CoV-2, we also included the proportion of individuals masking in public and NPI stringency as candidate predictors. After performing model selection of candidate models, parameter estimation of the final model was performed by restricted maximum likelihood.

## Acknowledgements

We thank the entire Seattle Flu Study (SFS) and Seattle Coronavirus Assessment Network (SCAN) team for their hard work and dedication to these projects and the study participants for their participation in this research. We also thank Public Health – Seattle & King County for their contributions to the SCAN study and for providing samples collected at King County COVID-19 drive-through testing sites, the Tacoma-Pierce County Health Department for permission to use SCAN samples collected in Pierce County, and the Rapid Health Information Network (RHINO) program at the Washington Department of Health for providing syndromic surveillance data. We thank Dr. Jeff Duchin for helpful comments on the manuscript and colleagues in the Fogarty International Center’s Division of International Epidemiology and Population Studies (DIEPS) and the Bedford Lab at Fred Hutch for useful discussions.

## Author contributions

Conceptualization: ACP, MF, JSh, TB, HYC, JAE, LMS, CV

Methodology: ACP, CH, RB, PDH, MF, JSh, TB, HYC, JAE, LMS, CV

Software: ACP, CH, RB, CM, DR, BC, MT, KSF, KK, BP, JL, TRS, JSt, AA, MF, CV

Validation: ACP, CH, RB, EM, LRS, LG, PDH, LMS

Formal analysis: ACP

Investigation: ACP

Resources: AA, MLJ, JSh, TB, HYC, JAE, LMS

Data acquisition or curation: ACP, CH, RB, CM, DR, BC, MT, KSF, KK, BP, ZA, JL, TRS, EM, LRS, JSt, LG, PDH, AA, AW, MLJ, MF, JSh, TB, HYC, JAE, LMS, CV

Writing—original draft: ACP

Writing—review & editing: ACP, CH, RB, SB, RP, MF, HYC, JAE, LMS, CV

Visualization: ACP

Supervision: SB, RP, KK, ZA, JSt, PDH, MF, JSh, TB, HYC, JAE, LMS, CV

Project administration: SB, RP, KK, ZA, JSt, PDH, MF, JSh, TB, HYC, JAE, LMS, CV

Funding acquisition: ACP, CH, SB, RP, AW, JSh, TB, HYC, JAE, LMS, CV

All authors have seen and approved the manuscript.

## Data and materials availability

Aggregated epidemiological and mobility data and code to reproduce analyses and figures will be made available in a GitHub repository and Dryad at the time of publication acceptance. Access to deidentified study participant data requires a signed data access agreement with the Seattle Flu Alliance and can be made available to researchers whose proposed use of the data is approved by study investigators. Requests for data access should be submitted to. Mobility metrics were generated using SafeGraph Weekly Patterns and Social Distancing datasets and Meta Data for Good Movement Range Maps, which were originally made freely available to academics in response to the COVID-19 pandemic. The SafeGraph Weekly Patterns dataset is currently available to academics for non-commercial use through an institutional university subscription or individual subscription to Dewey (https://www.deweydata.io/). The data access agreement with Dewey does not permit sharing of the raw data. Meta Data for Good Movement Range Maps are publicly accessible through the Humanitarian Data Exchange (https://data.humdata.org/dataset/movement-range-maps).

## Disclaimer

The findings and conclusions in this report are those of the authors and do not necessarily represent the official position of the US National Institutes of Health or the US government.

