## Supplementary Material for "Human mobility impacts the transmission of common respiratory viruses: A modeling study of the Seattle metropolitan area"

### Supplementary Methods

#### Seattle Flu Study surveillance arms

Recruitment sites and sample sizes are listed in Table S2.

***Community and clinic kiosks***. In the first year of SFS, participants with acute respiratory illness (ARI) were recruited at stand-alone kiosks in 7 clinical facilities (emergency departments, clinic and urgent care waiting rooms) and at 14 public sites, including the UW campus, SeaTac airport, workplaces, and high-traffic tourist areas. Participants were eligible to enroll if they had two or more new or worsening respiratory symptoms in the previous 7 days (fever, cough, sore throat, headache, diarrhea, nausea or vomiting, runny or stuffy nose, rash, fatigue, muscle or body aches, increased trouble with breathing, and/or ear pain or discharge) and were English- or Spanish-speaking. After completing a brief screening for eligibility to participate, individuals were consented. Upon enrolling, participants (or parent/guardian for minors) completed a questionnaire to collect participant demographics, illness characteristics, and behavioral and other clinical data. Trained research staff collected middle turbinate swabs for respiratory virus testing (Copan Diagnostics Inc., Murietta, CA). Participants received a $10 gift card for completing the study, and no additional study-related follow-up occurred. Participants were not permitted to re-enroll within a 7-day period.

***Outpatient clinics (Kaiser Permanente).*** From November 2018 to March 2020, participants seeking outpatient care for acute respiratory illness (ARI) at Seattle-based US Flu Vaccine Effectiveness (VE) Network sites were prospectively identified and recruited through Kaiser Permanente as part of the CDC Flu VE surveillance protocol (*1, 2*). Patients eligible for the CDC Flu VE study were aged at least 6 months of age and had a cough illness of < 8 days duration. Eligible and consenting patients (or parent/guardian for minors) were interviewed for demographics, risk factors for ARI, and influenza vaccination history. Study staff collected combined nasal and oropharyngeal swabs (nasal only in children aged < 2 years) for respiratory virus testing. In accordance with UW IRB approval, Health Insurance Portability and Accountability Act (HIPAA) authorization and written, informed consent was waived, as there was no direct contact with these participants or reasonable ability to recontact them for consent to participate in the study. Samples were obtained through a contractual agreement with Kaiser Permanente and transported to the study laboratory at the University of Washington (UW) for further molecular testing.

***Swab-and-Send Study.*** From October 2019 to March 2020, SFS deployed swab-and-send kits to collect nasal swabs from individuals in the community experiencing ARI. Study design, recruitment, and data collection are described in detail elsewhere (*3*). Briefly, study participants were recruited through referrals from health care providers, clinics, SFS community kiosks, schools, and workplaces, dissemination of printed flyers posted at community locations, and social media advertising. Individuals were eligible to participate in the study if they lived within the greater Seattle region, had experienced new or worsening cough and/or two ARI symptoms (fever, headache, sore throat or itchy/scratchy throat, nausea or vomiting, runny/stuffy nose or sneezing, fatigue, muscle or body aches, increased trouble with breathing, diarrhea, ear pain/discharge, or rash) within 7 days of enrollment, were English speaking, had a valid email address, and had access to the Internet at home. After an initial online screening questionnaire and consenting to participate in the research study, eligible participants completed an online enrollment questionnaire to provide their home address and contact information. Enrollees were mailed a home sample collection kit within 48 hours via private courier. Upon kit receipt, participants completed an online illness questionnaire to collect demographics, illness characteristics, and data on health behaviors. Samples were self-collected by participants 13 years and older via unsupervised middle turbinate swab (Copan Diagnostics Inc.). Parents or guardians performed swab collection for children younger than 13 years. Pediatric nasal swabs (Copan Diagnostics Inc.) were available for participants 5 years of age or younger. Participants were encouraged to return their nasal specimen within 24 hours or as soon as possible. Swab samples were returned to the study laboratory at UW via USPS Priority Mail prepaid postage, with a median time of 3 days from nasal swab collection to receipt at the lab (*3*).

***Greater Seattle Coronavirus Assessment Network.*** The greater Seattle Coronavirus Assessment Network (SCAN) was launched on March 23, 2020, and concluded in July 2022. Design, recruitment, and data collection for SCAN are described in detail elsewhere (*4, 5*). Briefly, SCAN was restricted to King County residents and recruited participants through social media advertising and community outreach. Eligibility criteria changed over time in response to testing demand and and were based on Public Use Microdata Areas (PUMA) and reported symptoms. Each PUMA had a daily allocation of enrollments, with over sampling of PUMAs in southern King County to ensure more equitable access to testing across the county population (*4, 5*). Study materials were available in English and 12 of the most spoken non-English languages in King County. Although symptom quotas changed over time, the majority of participants were symptomatic at the time of enrollment (> 90%)(*5*), with symptomatic enrollees defined as individuals who self-reported experiencing a new or worsening fever, cough, or shortness of breath within the past 7 days, and asymptomatic enrollees defined as individuals self-reporting none of these symptoms. In addition to community enrollments, some participants were invited as part of PHSKC's contact tracing efforts or through collaborations with community-based organizations to increase testing of underrepresented or high-risk populations; these samples were excluded from the analysis. After an initial online screening questionnaire, eligible participants (or parent/guardian for minors) were prompted to complete a detailed demographic and health behavior questionnaire. Within 24 hours of enrollment, sample collection kits were delivered via private courier. Samples were self-collected by participants aged 13 years and older via unsupervised middle turbinate or anterior nares swabs. Parents or guardians performed swab collection for children younger than 13 years of age. Swab samples were picked up by private courier on the morning after delivery and returned within 24 hours to the study laboratory at UW for testing.

***King County COVID-19 drive-through testing sites.*** Beginning in April 2021, SFS obtained residual nasal swab specimens collected at eight Public Health Seattle-King County (PHSKC) COVID-19 drive-through testing sites. In accordance with UW IRB approval, HIPAA authorization and written, informed consent was waived, as there was no direct contact with these participants or reasonable ability to recontact them for consent to participate in the study. Samples were obtained through a contractual agreement with UW Virology, which conducted the SARS-CoV-2 testing for PHSKC drive-through sites. At the time of testing, individuals completed an optional questionnaire that collected demographics, the reason for testing, COVID-19 vaccination status, whether they are currently symptomatic, and, if symptomatic, the number of days since symptom onset. Samples from both symptomatic and asymptomatic individuals were obtained by SFS, with symptomatic individuals defined as those who answered "yes" to the question "Do you have COVID-19 symptoms now?"

***Residual hospital samples.*** Since the inception of the study, SFS obtained residual nasal swab specimens collected at clinician discretion from major hospitals in the Seattle area, including Seattle Children's, UW Medical Center, Northwest Hospital, and Harborview Medical Center. In April 2020, surveillance from UW Medical Center and Northwest Hospital discontinued. Samples were linked with demographic and clinical metadata extracted from the patients' electronic medical records (EMR). In accordance with UW IRB approval, HIPAA authorization and written, informed consent was waived, as there was no direct contact with these participants or reasonable ability to recontact them for consent to participate in the study. Samples were obtained through contractual agreements with each medical center and transported to the study laboratory at UW for further molecular testing. Encounter IDs and medical record numbers (MRNs) in combination were used as unique patient identifiers at sites except Seattle Children's, where a unique patient ID was created. Prior to March 2020, most hospital residuals were collected from patients experiencing ARI. After March 2020, there was increased testing of asymptomatic individuals at hospitals due to pre-procedure or surveillance testing for COVID-19. We used ICD-10 codes specific to respiratory illness (Harborview Medical Center, Northwest Hospital, and UW Medical Center) (Table S5) or pre-procedure COVID-19 testing flags (Seattle Children's) to distinguish symptomatic and asymptomatic patients.

**Statistical Analysis**

***Short-term forecasting of daily transmissibility using mobility data.*** We built predictive models of daily respiratory virus transmission, using cell phone mobility trends, the co-circulation of other respiratory viruses, and activity of the target virus during the previous two weeks (14 autoregressive terms) as input variables. Similar to an approach for forecasting influenza-like illness activity (AutoRegression with GOogle search data)(*6*), our models implemented L1 regularization (LASSO) to automatically select the most relevant terms for predicting Rt up to 7-days ahead, using a moving window for the training period (that immediately precedes the dates of estimation) to capture the most recent changes in mobility behavior and viral activity (*6*). To better observe changes in predictive performance and variable selection over time, we focused on three respiratory viruses that circulated continuously throughout most of the study period: hRV, AdV, and SARS-CoV-2. In addition to mobility and autoregressive terms, we included proxies for viral-viral interactions, wherein SARS-CoV-2 Rt was a covariate in the hRV and AdV models, and hRV Rt was a covariate in the SARS-CoV-2 model. We also tested models that included daily precipitation, average wet bulb temperature, and average relative humidity as covariates.

We compared each full model's estimates, along with those of reduced models including only AR terms, only mobility terms, or only mobility and pathogen interaction terms, to observed Rt values by calculating several accuracy metrics, including root-mean-squared error (RMSE), mean absolute error (MAE), mean absolute percentage error (MAPE), and the Pearson correlation between observed and predicted Rt (Table S2). For all three viruses, we found that one-month moving windows produced the most accurate forecasts of Rt, though there were few discernible trends in which mobility terms were retained over time. Expanding the training window produced clearer patterns of which mobility terms were consistently retained by the model but at the expense of predictive accuracy.

### Supplementary Results

#### Real-time tracking of rhinovirus, adenovirus, and SARS-CoV-2 transmission

For all three viruses, full models with mobility terms, pathogen interaction terms, and autoregressive (AR) terms and models with only autoregressive (AR) terms had similar predictive accuracy and outperformed models with only mobility terms or only mobility and pathogen interaction terms (Table S2); thus, tracking mobility behavior is not essential for predicting respiratory virus transmission. However, mobility-only models still produced accurate forecasts across the entire study period (Pearson's r with observed data, hRV: 0.92; AdV: 0.82; SARS-CoV-2: 0.83) (Table S2). We also tested models incorporating local temperature, precipitation, and absolute humidity but climatic variables did not improve model performance, potentially because non-enveloped viruses and pandemic SARS-CoV-2 do not exhibit strong seasonality.

Among candidate mobility predictors, the percentage of devices leaving home, between-neighborhood movement, and the inflow of outside visitors had the highest mean (absolute) coefficient values and were the most frequently retained variables across moving training windows. To determine if mobility data are more useful in predicting Rt during drastic changes in population movement, we calculated accuracy metrics for the period including Seattle's stay-at-home orders and the initial lifting of restrictions (February 28 – June 30, 2020). In terms of RMSE, models with only mobility terms were 48% (AdV), 20% (hRV), and 19% (SARS-CoV-2) more accurate during the first half of 2020 compared to the entire study period (Table S2). Thus, monitoring major changes in mobility could be helpful for general situational awareness and planning purposes.

### Supplementary Figures

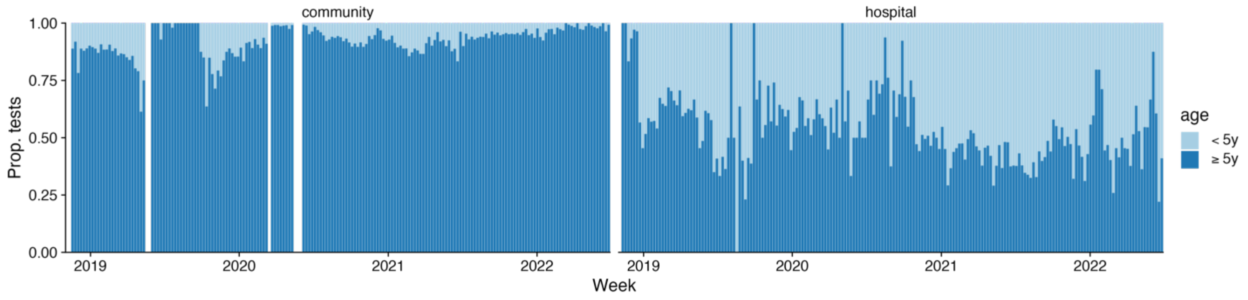


**Figure S1. The weekly age distribution of respiratory specimens collected from community settings and hospitals, November 2018 – June 2022.** Bars are colored by the proportion of samples collected from individuals aged <5 (light blue) or ≥5 years (dark blue). Sample sources for community-based testing include swab-and-send at-home testing programs, kiosks in high foot traffic areas, outpatient clinics, and King County COVID-19 drive-through testing sites.

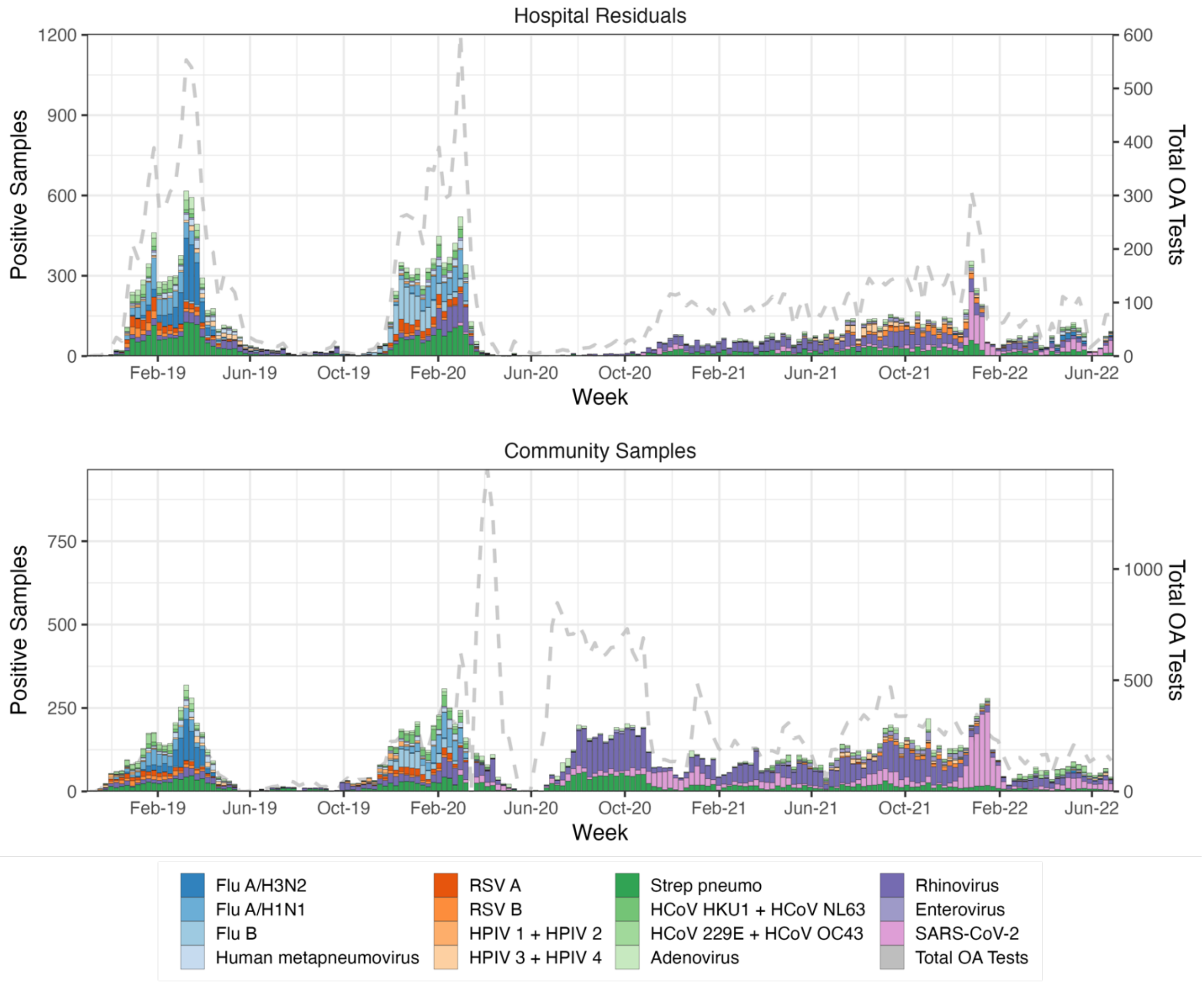


**Figure S2. The weekly number of samples testing positive for respiratory pathogens in hospitals (top) and community settings (bottom).** Colored bars represent the number of samples testing positive for each pathogen. The gray dashed line is the number of respiratory specimens tested on the OpenArray (OA) platform. The left y-axis corresponds to the number of positive samples collected each week, and the right y-axis corresponds to the total number of specimens collected each week and tested on OA. Sources for community-based samples (bottom) include swab-and-send at-home testing programs, kiosks in high foot traffic areas, outpatient clinics, and King County COVID-19 drive-through testing sites.

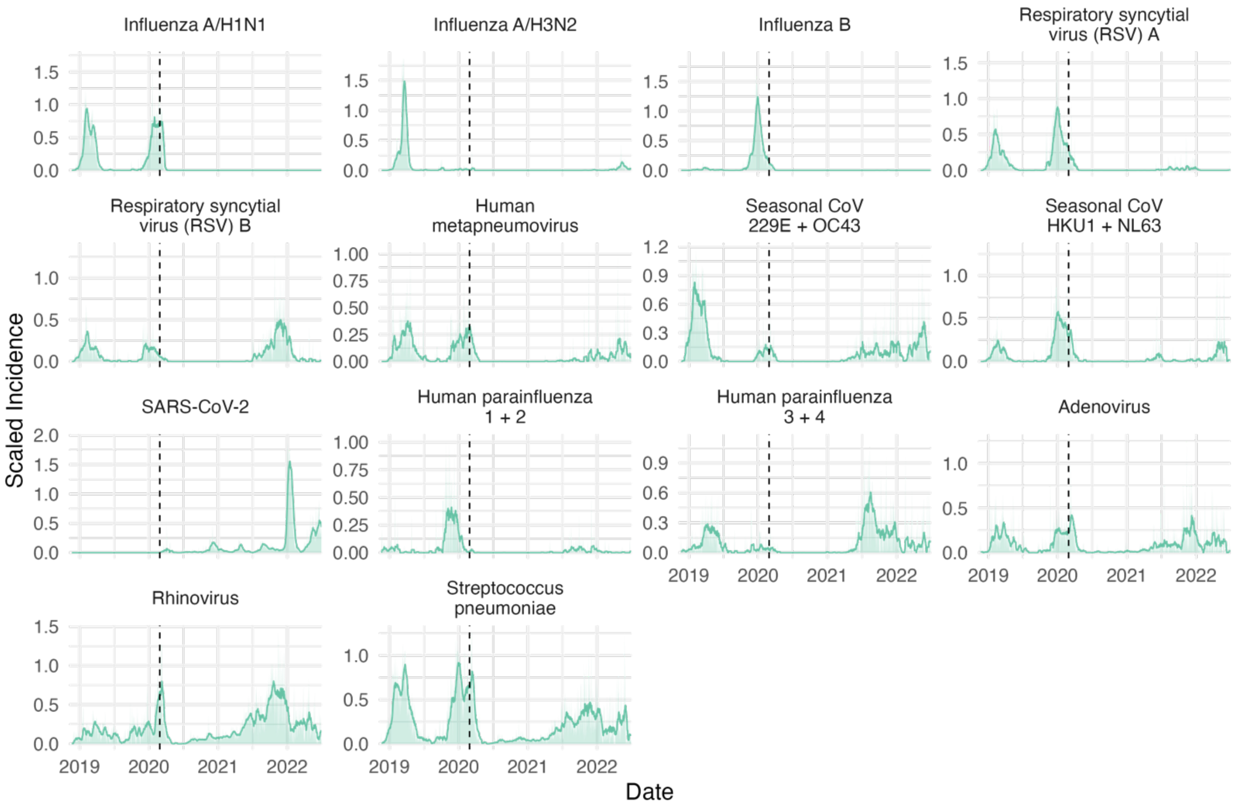


**Figure S3. Reconstructed daily incidences of individual respiratory pathogens, adjusted for testing volume over time, age, clinical setting, and local syndromic respiratory illness rates.** Hospital and community-based incidences were rescaled to fall between 0 and 1 and summed to aid in comparing relative changes in incidence between pathogens over time. We applied two-week rolling averages to incidences to reduce noise. The vertical dashed line indicates the date of Washington's State of Emergency declaration (February 29, 2020).

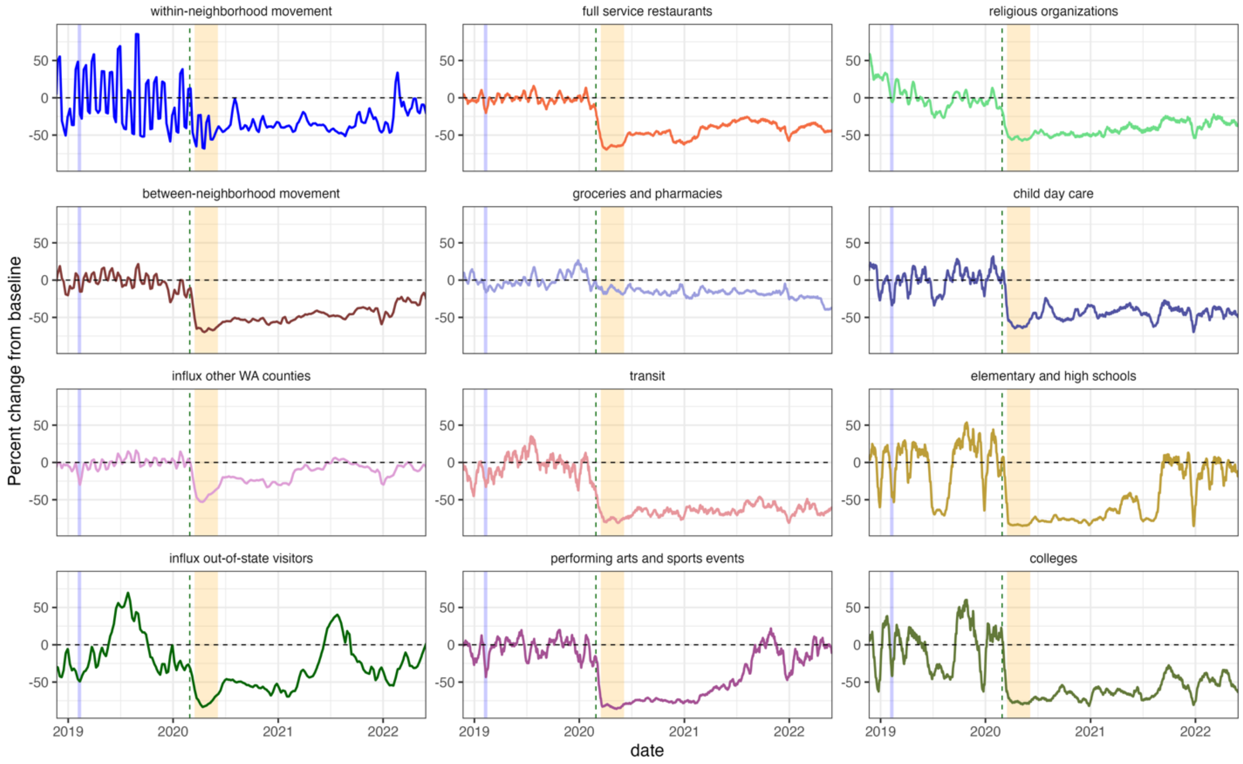


**Figure S4. Cell phone mobility metrics for Seattle-King County, Washington, November 2018 – June 2022.** For each mobility indicator, we summed daily or weekly visits for each point of interest (POI) category and measured the percent change in movement over time relative to the average movement observed in all of 2019 (excluding national holidays) and applied a two-week rolling average to reduce noise. The vertical blue shaded panel indicates the timing of a major snowstorm in Seattle (February 3-15, 2019), the vertical dashed line indicates the date of Washington's State of Emergency declaration (February 29, 2020), and the vertical orange shaded panel indicates Seattle's stay-at-home period (March 23 – June 5, 2020).

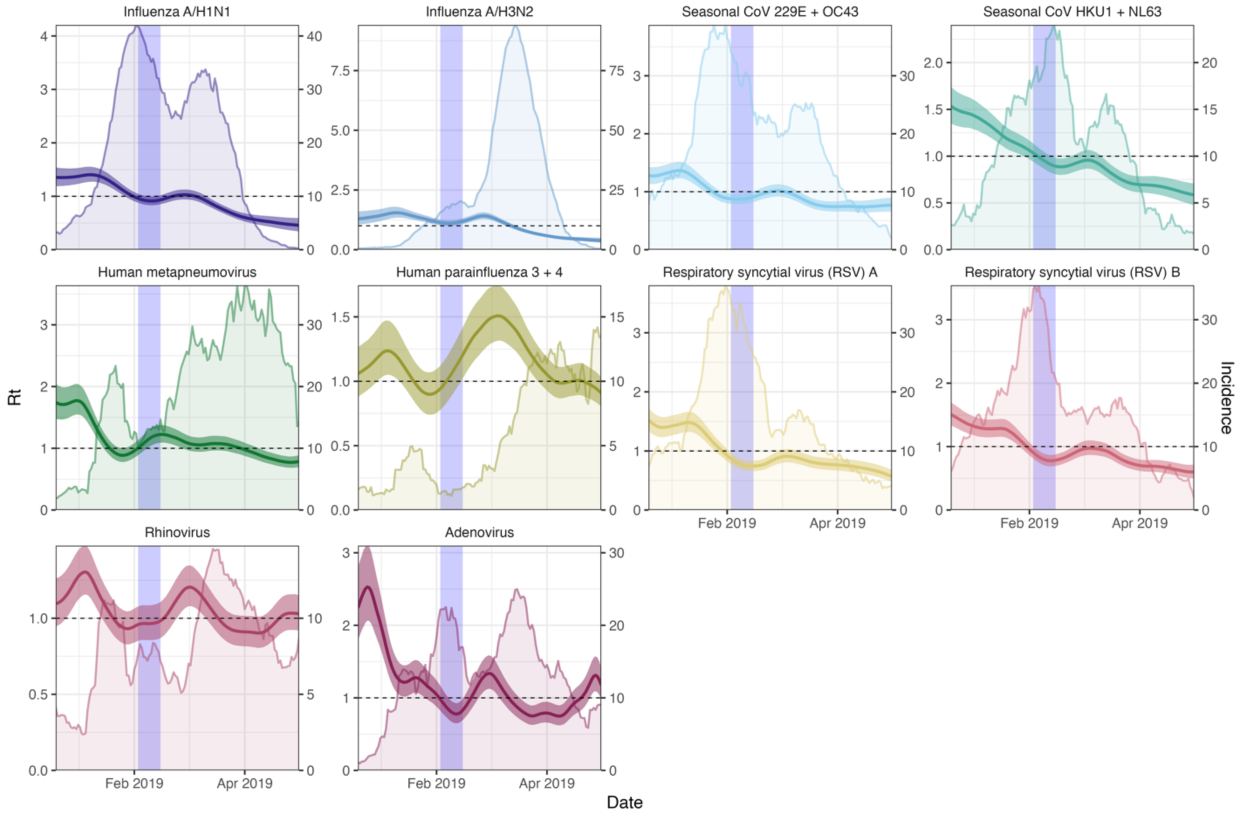


**Figure S5. Daily incidences and transmissibility of respiratory viruses circulating in the greater Seattle region, December 2018 – May 2019.** Daily time-varying effective reproduction numbers (Rt, thick lines, left y-axis) and reconstructed incidences of respiratory viruses (thin lines, right y-axis). We applied two-week rolling averages to incidences to reduce noise. The vertical blue shaded panel indicates the timing of a major snowstorm (February 3 – 15, 2019).

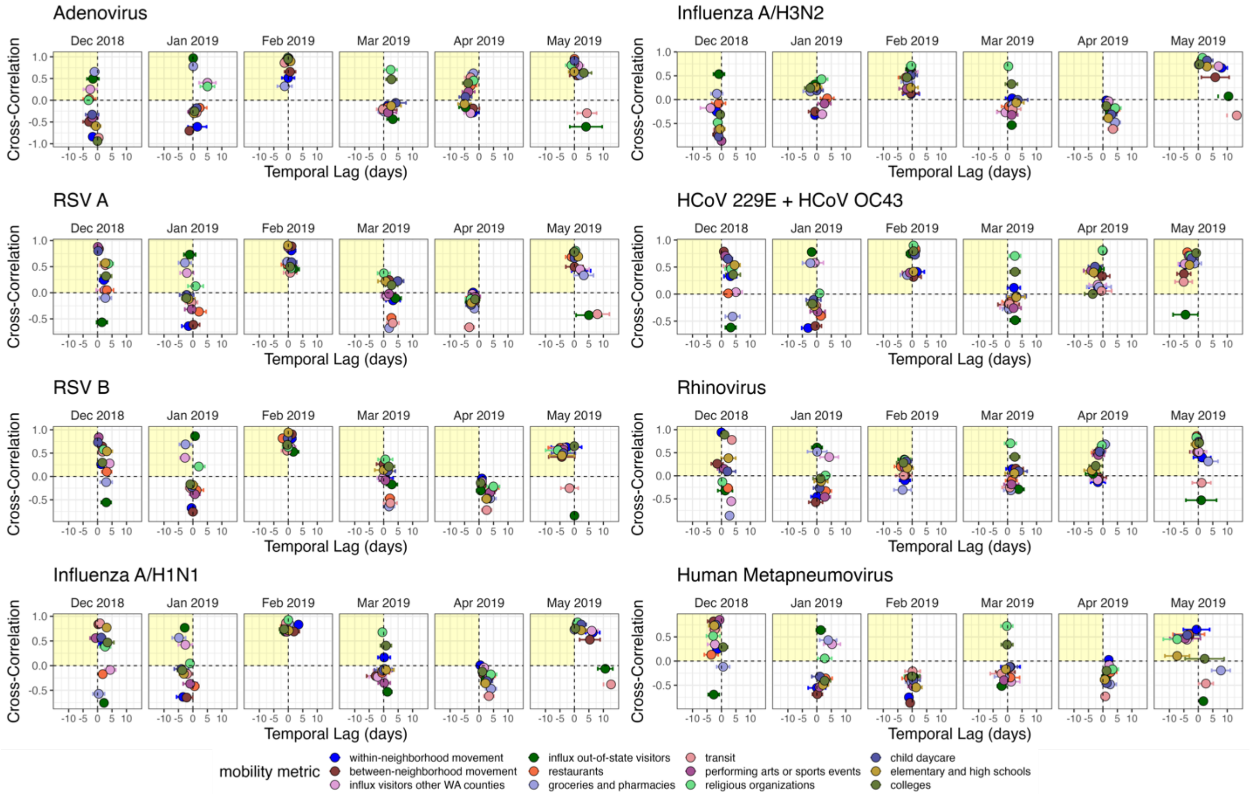


**Figure S6. Time series cross-correlations and optimal lags between respiratory pathogen transmissibility (daily effective reproduction numbers, Rt) and cell phone mobility in the greater Seattle region, December 2018 – May 2019**. Weekly time series cross correlations in moving 5-month windows were averaged by calendar month. Points are individual mobility indicators derived from SafeGraph mobile device location data. Correlation coefficients are shown on the y-axis, and temporal lags (in weeks) between Rt and mobility are shown on the x-axis. Negative temporal lags indicate behavior leads Rt, and positive temporal lags indicate Rt leads behavior. The yellow shaded panel in each facet includes mobility indicators that have a leading, positive relationship with transmission, and hence would be considered predictive of transmission.

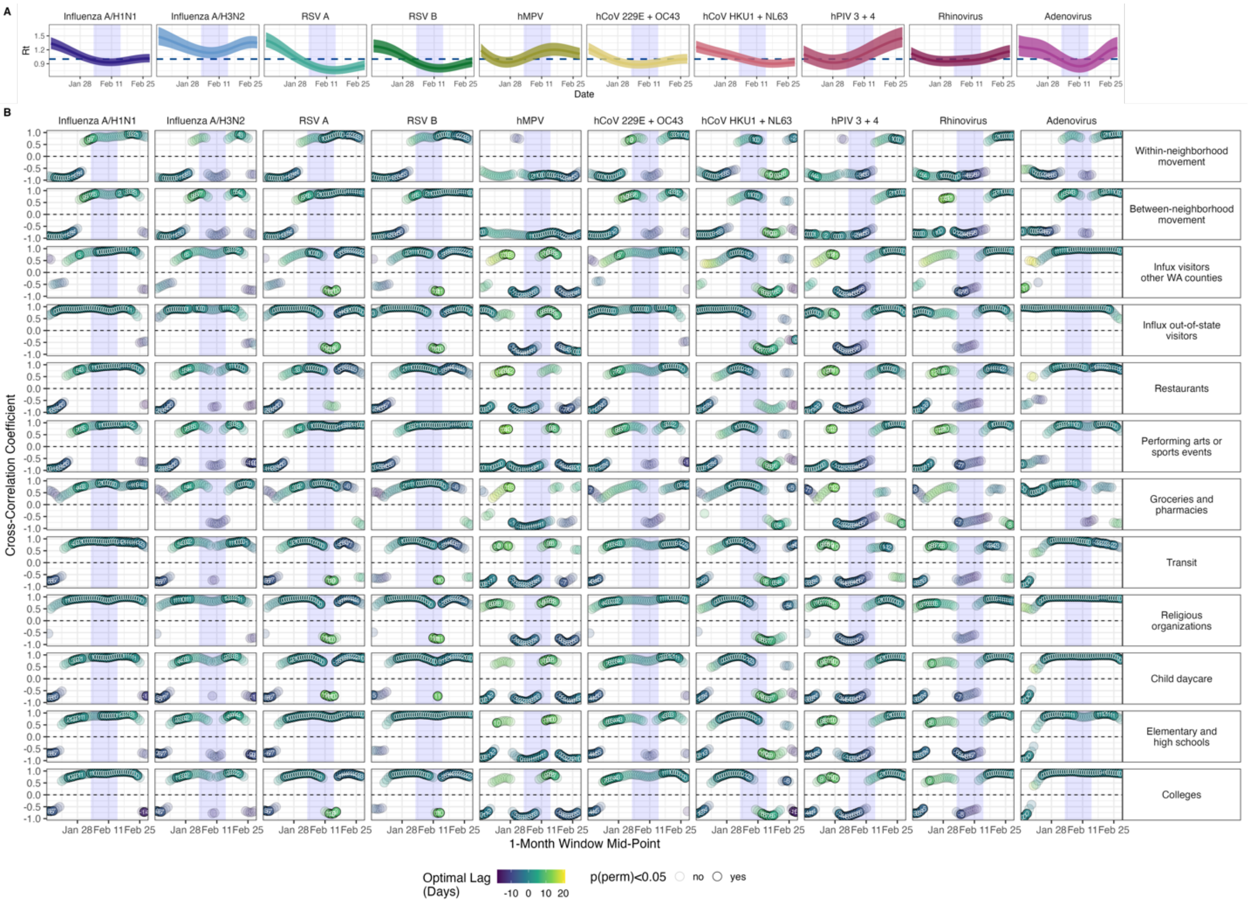


**Figure S7. A. Daily effective reproduction numbers (Rt) of respiratory viruses circulating in the greater Seattle region, and B. Rolling daily cross-correlations between pathogen transmissibility and cell phone mobility during January – February 2019.** Points represent the maximum coefficient values for 1-month rolling cross-correlations between daily effective reproduction numbers (Rt) and individual mobility metrics. Point color and the number within each point indicate the lag in weeks corresponding to the maximum cross-correlation coefficient value for each 1-month period ("optimal lag"). Negative values indicate that mobility leads Rt, and positive values indicate that mobility lags Rt. A lag of 0 indicates that the time series are in phase. Point transparency indicates statistical significance based on 1000 block bootstrap permutations (yes: solid, no: transparent). The vertical blue shaded panel indicates the timing of a major snowstorm (February 3 – 15, 2019).

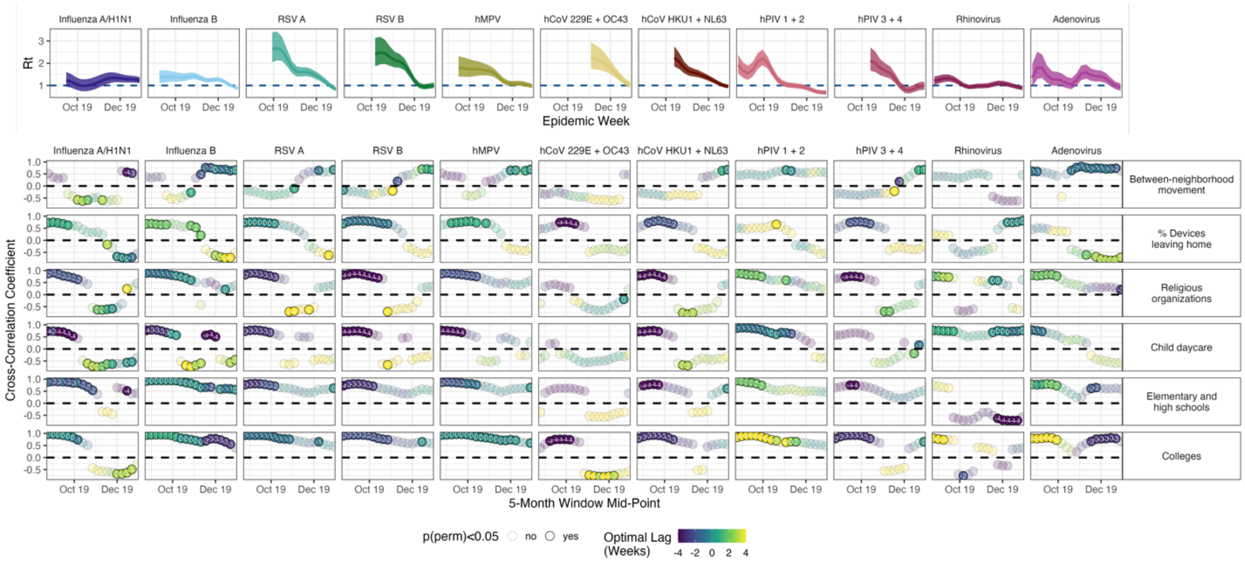


**Figure S8. A. Weekly effective reproduction numbers (Rt) of respiratory viruses circulating in the greater Seattle region, and B. Rolling cross-correlations between pathogen transmissibility and cell phone mobility during the 2019-2020 winter season prior to the start of the COVID-19 pandemic, August 2019 – January 2020.** Points represent the maximum coefficient values for 5-month rolling cross-correlations between weekly effective reproduction numbers (Rt) and individual mobility metrics. Point color and the number within each point indicate the lag in weeks corresponding to the maximum cross-correlation coefficient value for each 5-month period ("optimal lag"). Negative values indicate that mobility leads Rt, and positive values indicate that mobility lags Rt. A lag of 0 indicates that the time series are in phase. Point transparency indicates statistical significance based on 1000 block bootstrap permutations (yes: solid, no: transparent).

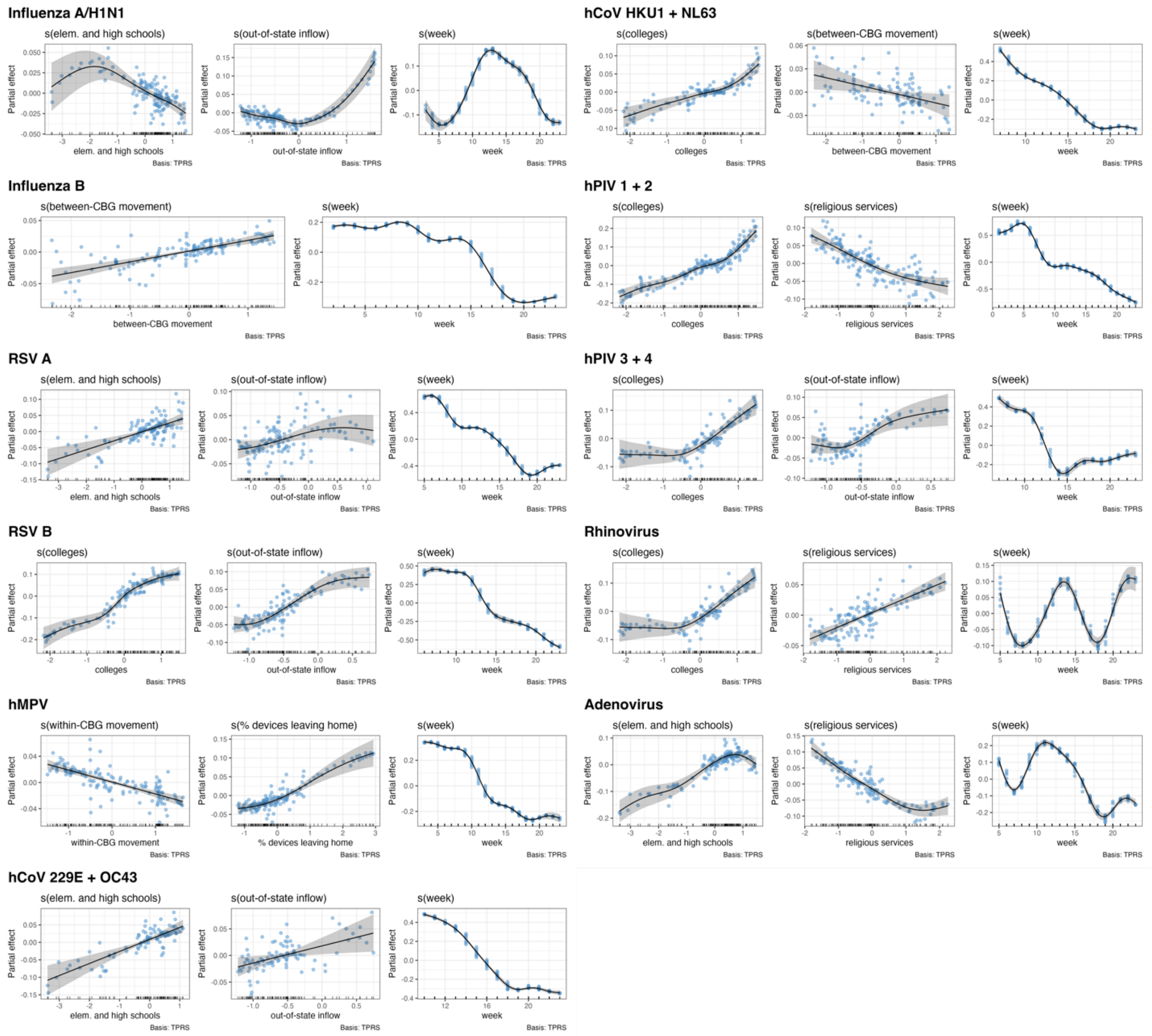


**Figure S9. Generalized additive model (GAM) plots showing the partial effects of selected mobility indicators and time trends on the daily effective reproduction numbers (Rt) of endemic respiratory viruses during the 2019-2020 winter season prior to the start of the COVID-19 pandemic, September 2019 – January 2020.** Tick marks on the x-axis are observed data points. The y-axis represents of the partial effect of each variable. Shaded areas indicate the 95% confidence intervals of partial effects. The blue points are partial residuals.

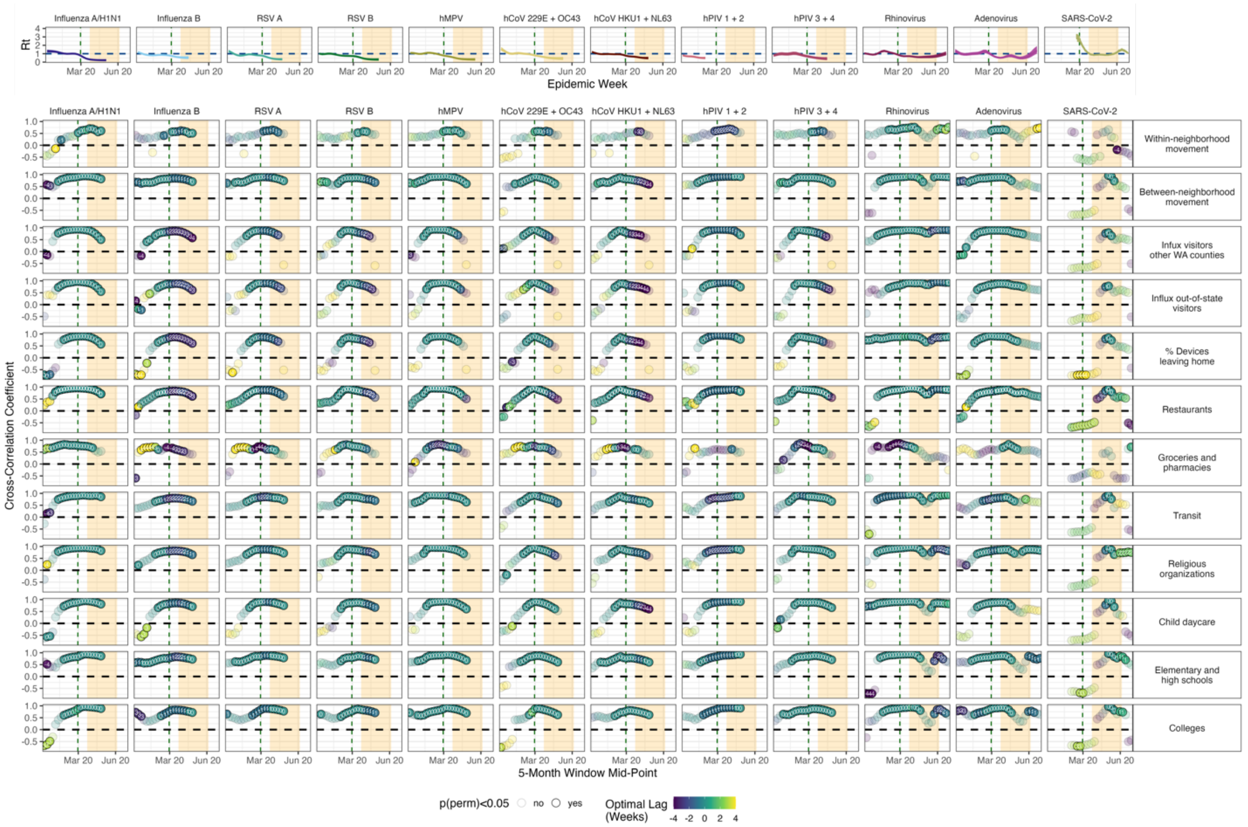


**Figure S10. A. Weekly effective reproduction numbers (Rt) of respiratory viruses circulating in the greater Seattle region, and B. Rolling cross-correlations between pathogen transmissibility and cell phone mobility during the initial months of the COVID-19 pandemic, December 2019 – June 2020.** Points represent the maximum coefficient values for 5-month rolling cross-correlations between weekly effective reproduction numbers (Rt) and individual mobility metrics. Point color and the number within each point indicate the lag in weeks corresponding to the maximum cross-correlation coefficient value for each 5-month period ("optimal lag"). Negative values indicate that mobility leads Rt, and positive values indicate that mobility lags Rt. A lag of 0 indicates that the time series are in phase. Point transparency indicates statistical significance based on 1000 block bootstrap permutations (yes: solid, no: transparent). The vertical dashed line indicates the date of Washington's State of Emergency declaration (February 29, 2020), and the vertical orange shaded panel indicates Seattle's stay-at-home period (March 23 – June 5, 2020).

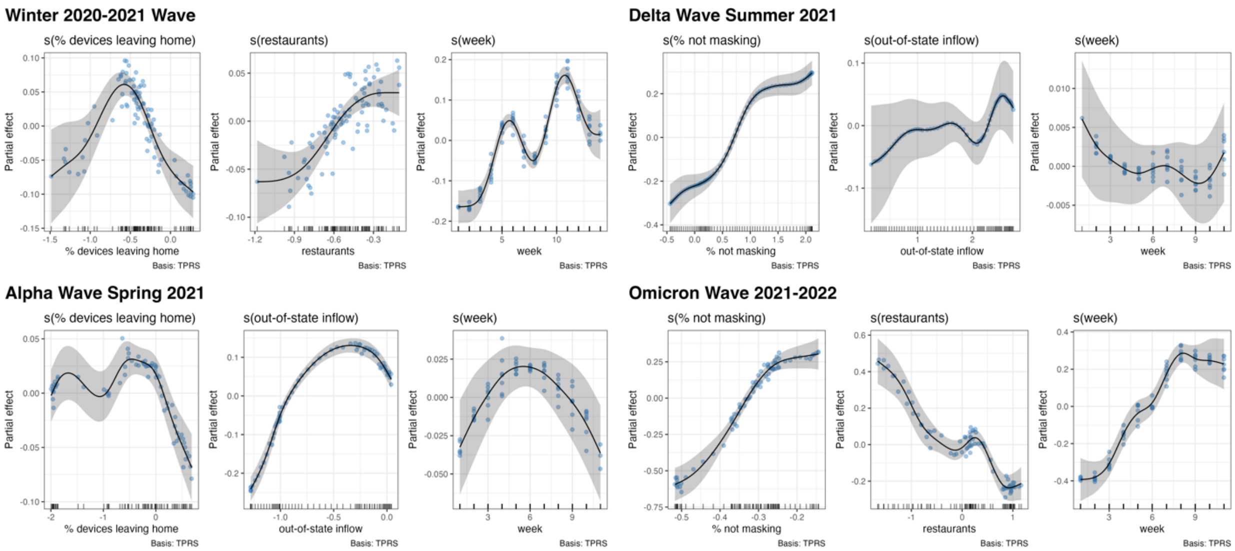


**Figure S11. Generalized additive model (GAM) plots showing the partial effects of selected mobility indicators and time trends on the daily effective reproduction numbers (Rt) of SARS-CoV-2 during four COVID-19 waves in Seattle: the winter 2020-2021 wave, the Alpha wave in Spring 2021, the Delta Wave in Summer 2021, and the Omicron BA.1 wave during late 2021 to early 2022.** Tick marks on the x-axis are observed data points. The y-axis represents of the partial effect of each variable. Shaded areas indicate the 95% confidence intervals of partial effects. The blue points are partial residuals.

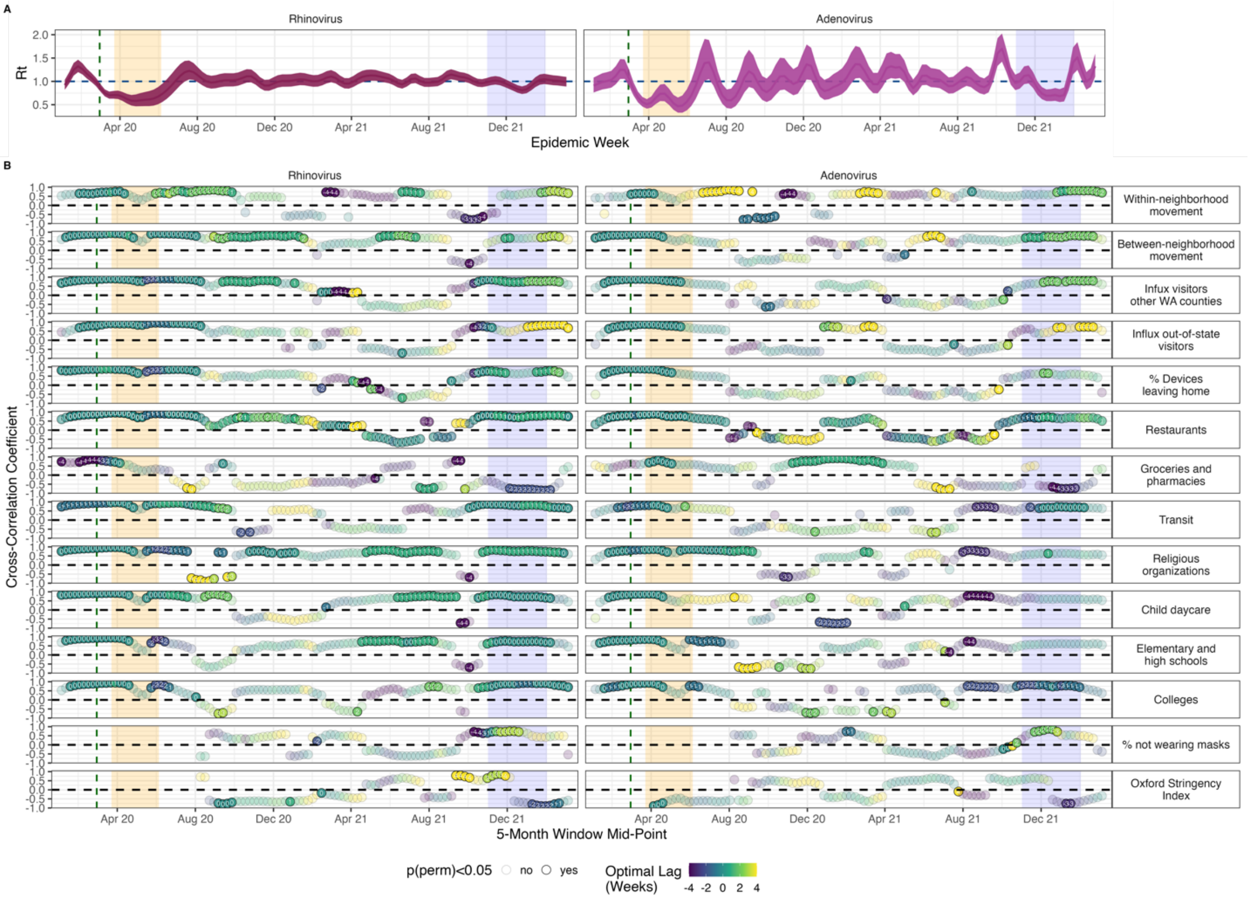


**Figure S12. A. Weekly effective reproduction numbers (Rt) of two non-enveloped viruses – rhinovirus and adenovirus – circulating in the greater Seattle region, and B. Rolling cross-correlations between pathogen transmissibility and cell phone mobility during the COVID-19 pandemic, January 2020 – March 2022.** Points represent the maximum coefficient values for 5-month rolling cross-correlations between weekly effective reproduction numbers (Rt) and individual behavioral metrics. Point color and the number within each point indicate the lag in weeks corresponding to the maximum cross-correlation coefficient value for each 5-month period (“optimal lag”). Negative values indicate that behavior leads Rt, and positive values indicate that behavior lags Rt. A lag of 0 indicates that the time series are in phase. Point transparency indicates statistical significance based on 1000 block bootstrap permutations (yes: solid, no: transparent). The vertical dashed line indicates the date of Washington’s State of Emergency declaration (February 29, 2020), the vertical orange shaded panel indicates Seattle’s stay-at-home period (March 23 – June 5, 2020), and the vertical blue shaded panel indicates the timing of the Omicron BA.1 wave (November 2021 – January 2022).

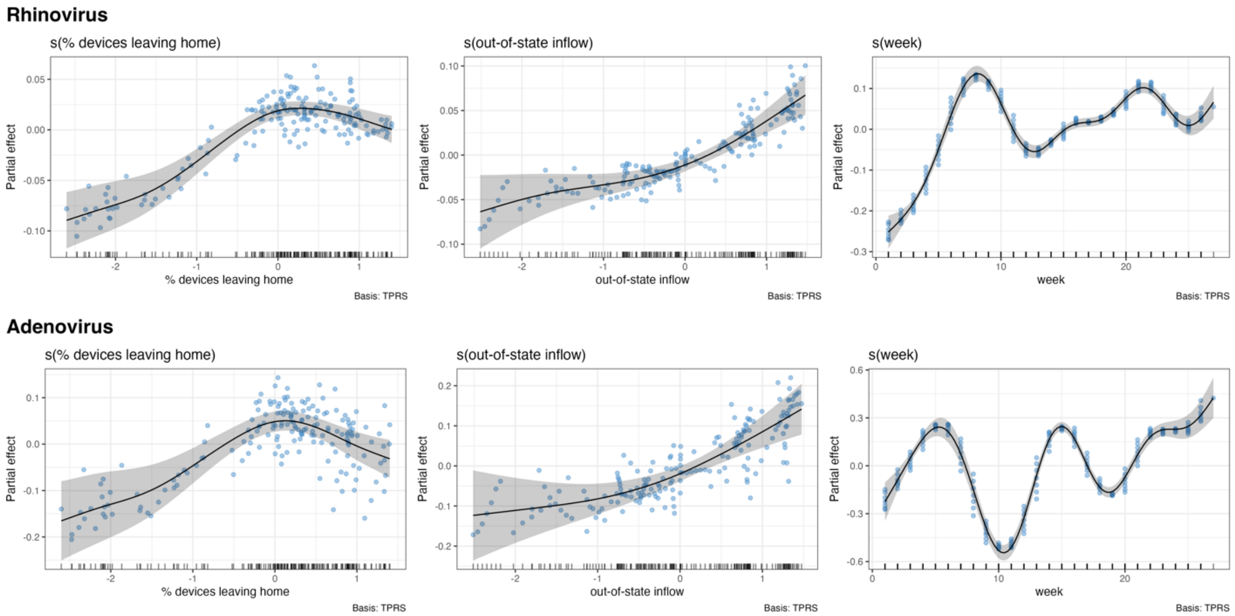


**Figure S13. Generalized additive model (GAM) plots showing the partial effects of selected mobility indicators and time trends on the daily effective reproduction numbers (Rt) of two non-enveloped respiratory viruses – rhinovirus and adenovirus – during their first six months of rebound, June 2020 – November 2020.** Tick marks on the x-axis are observed data points. The y-axis represents of the partial effect of each variable. Shaded areas indicate the 95% confidence intervals of partial effects. The blue points are partial residuals.

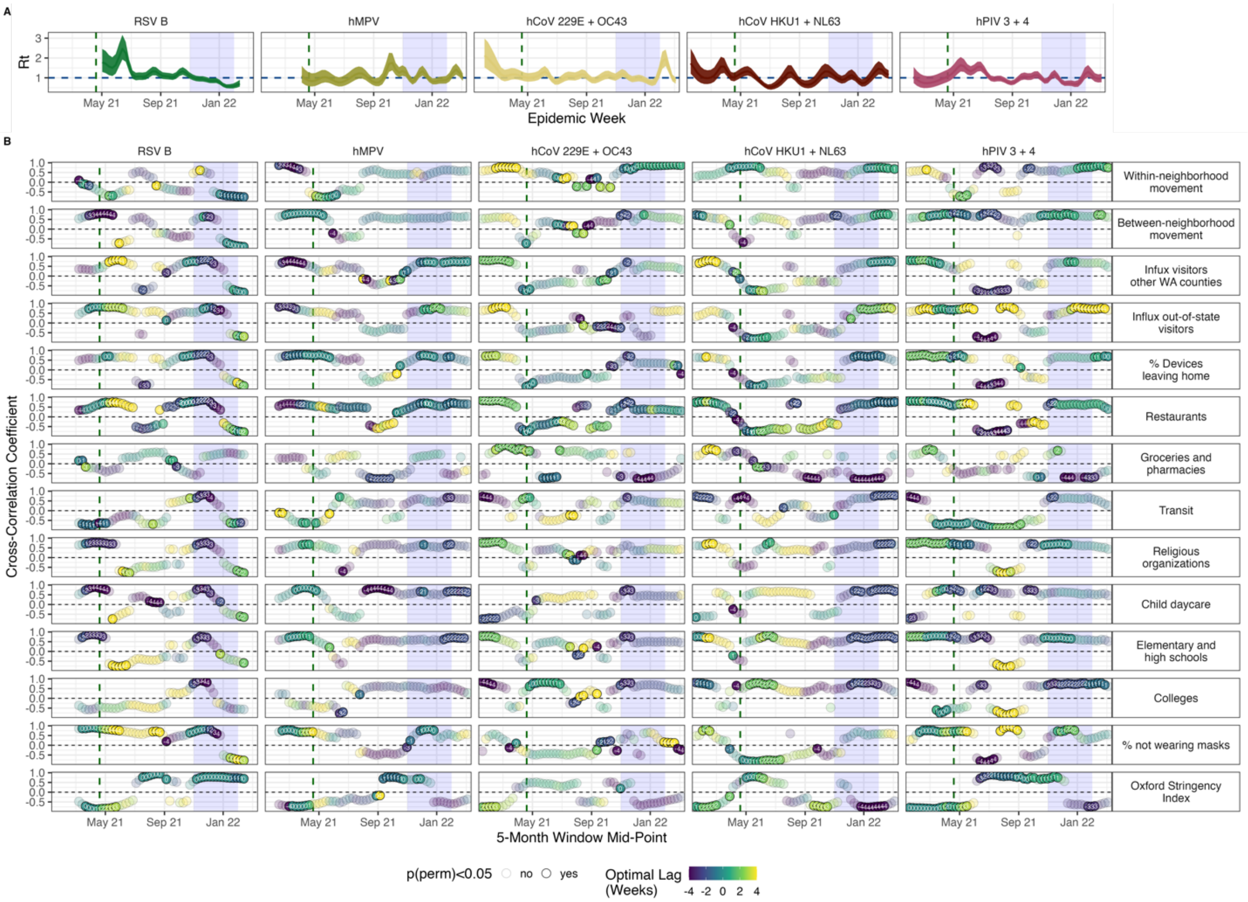


**Figure S14. A. Weekly effective reproduction numbers (Rt) of enveloped viruses circulating in the greater Seattle, WA, region, and B. Rolling cross-correlations between pathogen transmissibility and cell phone mobility during the COVID-19 pandemic, January 2021 – March 2022.** Points represent the maximum coefficient values for 5-month rolling cross-correlations between weekly effective reproduction numbers (Rt) and individual mobility and behavioral metrics. Point color and the number within each point indicate the lag in weeks corresponding to the maximum cross-correlation coefficient value for each 5-month period ("optimal lag"). Negative values indicate that behavior leads Rt, and positive values indicate that behavior lags Rt. A lag of 0 indicates that the time series are in phase. Point transparency indicates statistical significance based on 1000 block bootstrap permutations (yes: solid, no: transparent). The vertical dashed line indicates when Washington public schools were required to offer at least two days of partial in-person instruction to all grades (April 19, 2021), and the vertical blue shaded panel indicates the timing of the Omicron BA.1 wave (November 2021 – January 2022).

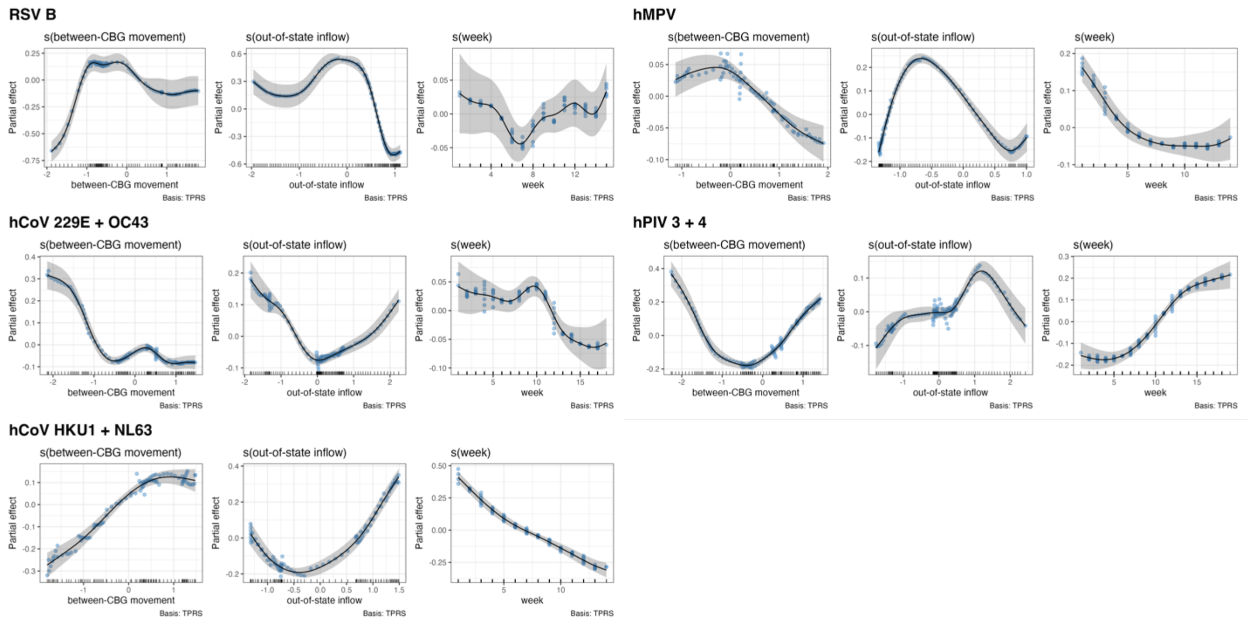


**Figure S15. Generalized additive model (GAM) plots showing the partial effects of selected mobility indicators and time trends on the daily effective reproduction numbers (Rt) of enveloped respiratory viruses during their initial months of rebound, January – August 2021.** Tick marks on the x-axis are observed data points. The y-axis represents of the partial effect of each variable. Shaded areas indicate the 95% confidence intervals of partial effects. The blue points are partial residuals.

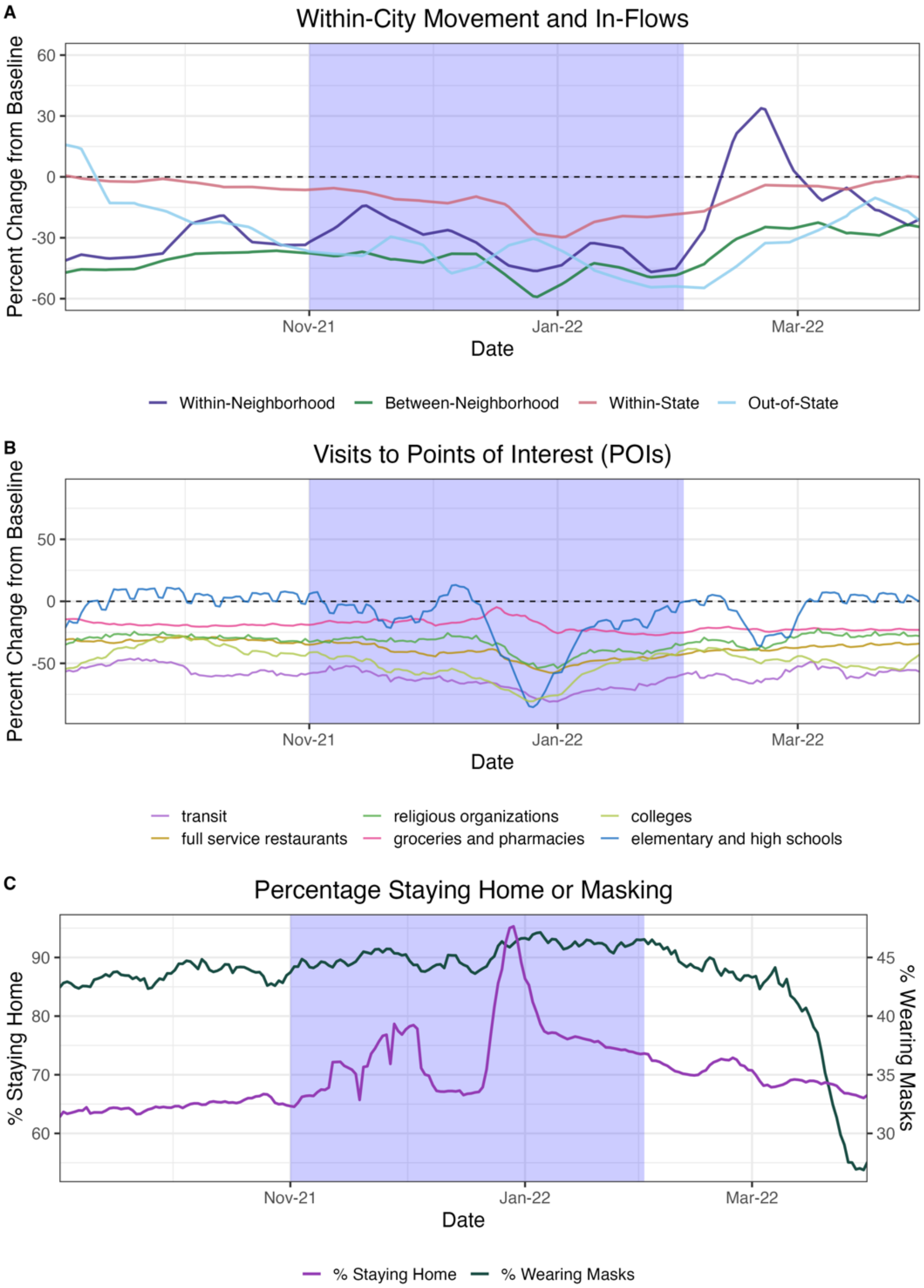


**Figure S16. Mobility and masking trends in the greater Seattle, Washington region during November 2021 – April 2022 based on cell phone location.** In each panel, the vertical blue shaded panel indicates the timing of the Omicron BA.1 wave in Seattle (November 2021 – February 2022). **A.** The percent change from baseline for large-scale population movements: inflow of out-of-state visitors, inflow of visitors from other WA counties, between-neighborhood movement of King County residents, and within-neighborhood movement of King County residents. **B.** The percent change from baseline in foot traffic to different categories of points of interest (POIs): transit stations, religious organizations, colleges and universities, full-service restaurants, groceries and pharmacies, and elementary and secondary schools. **C.** The percentage of devices staying completely at home (purple) and the percentage of King County residents masking in public (dark green).

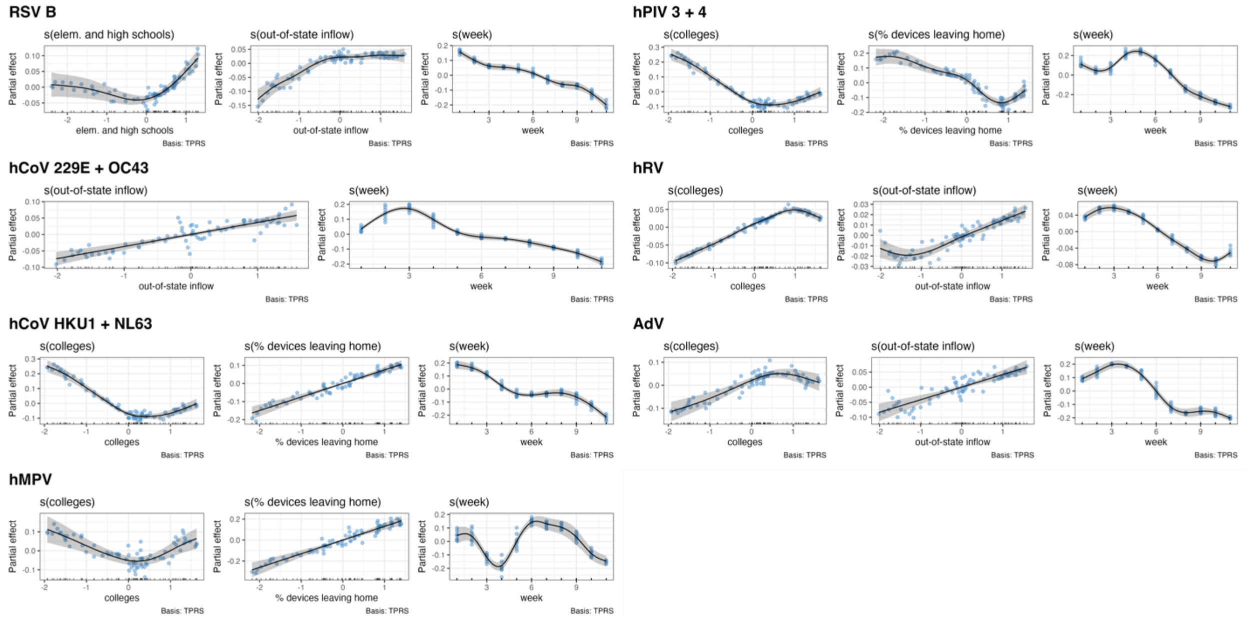


**Figure S17. Generalized additive model (GAM) plots showing the partial effects of selected mobility indicators and time trends on the daily effective reproduction numbers of endemic respiratory viruses during the beginning of the Omicron BA.1 wave, November 2021 – January 2022.** Tick marks on the x-axis are observed data points. The y-axis represents of the partial effect of each variable. Shaded areas indicate the 95% confidence intervals of partial effects. The blue points are partial residuals.

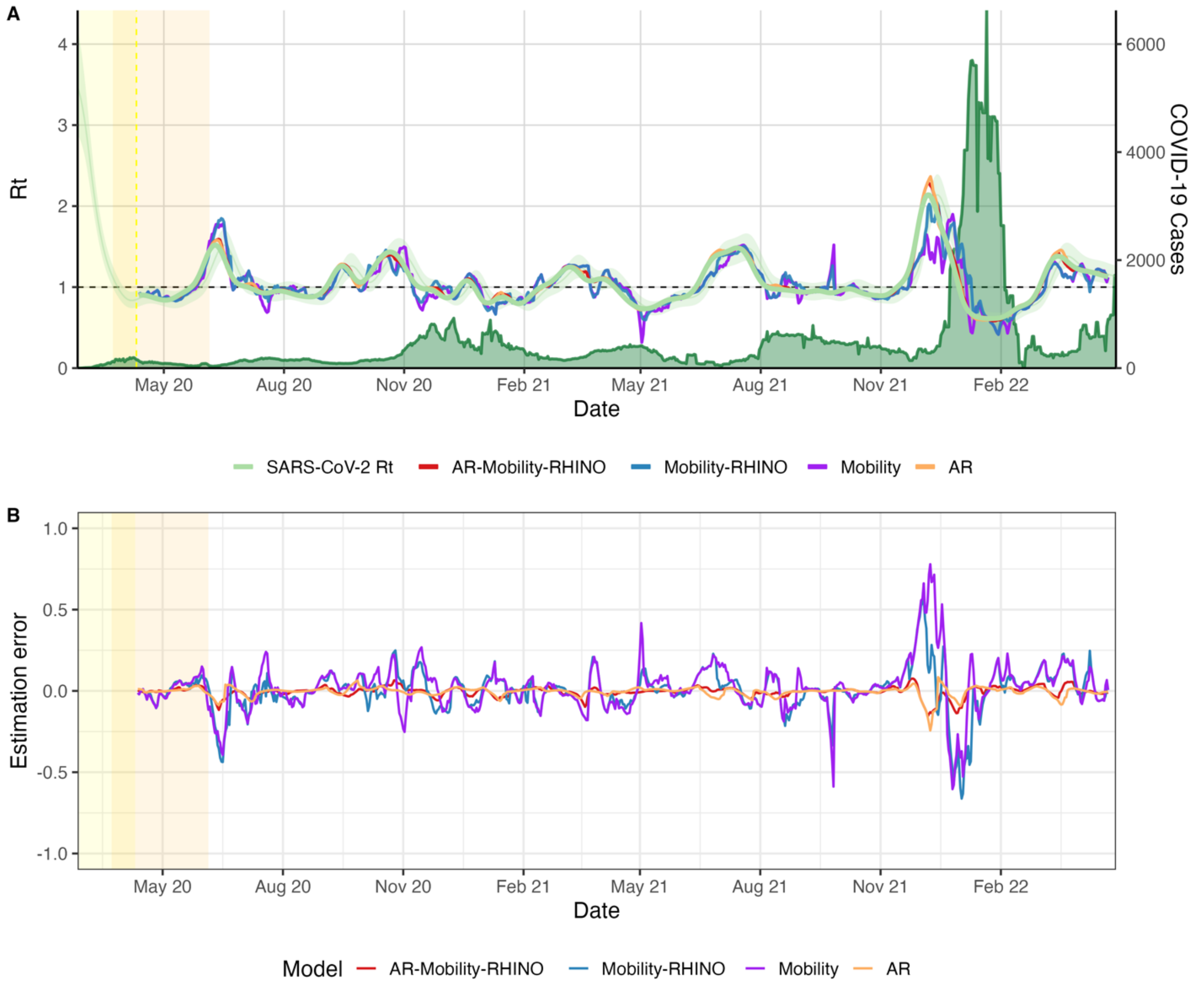


**Figure S18. Predictive model of daily SARS-CoV-2 transmission (time-varying effective reproduction number, Rt).** The yellow shaded area indicates the initial model training period, and the orange shaded area indicates Seattle's stay-at-home period. **A.** Estimated daily transmissibility (Rt) (left y-axis) from the full model with autoregressive (AR) terms, mobility terms, and rhinovirus (hRV) interaction terms (red), contrasting with observed Rt (light green) and estimates from a model with only mobility and hRV interaction terms (blue), a model with only mobility terms (purple), and a model with only AR terms (orange). Daily COVID-19 cases are shaded dark green (right y-axis). **B.** Model estimation error, defined as observed Rt minus predicted Rt.

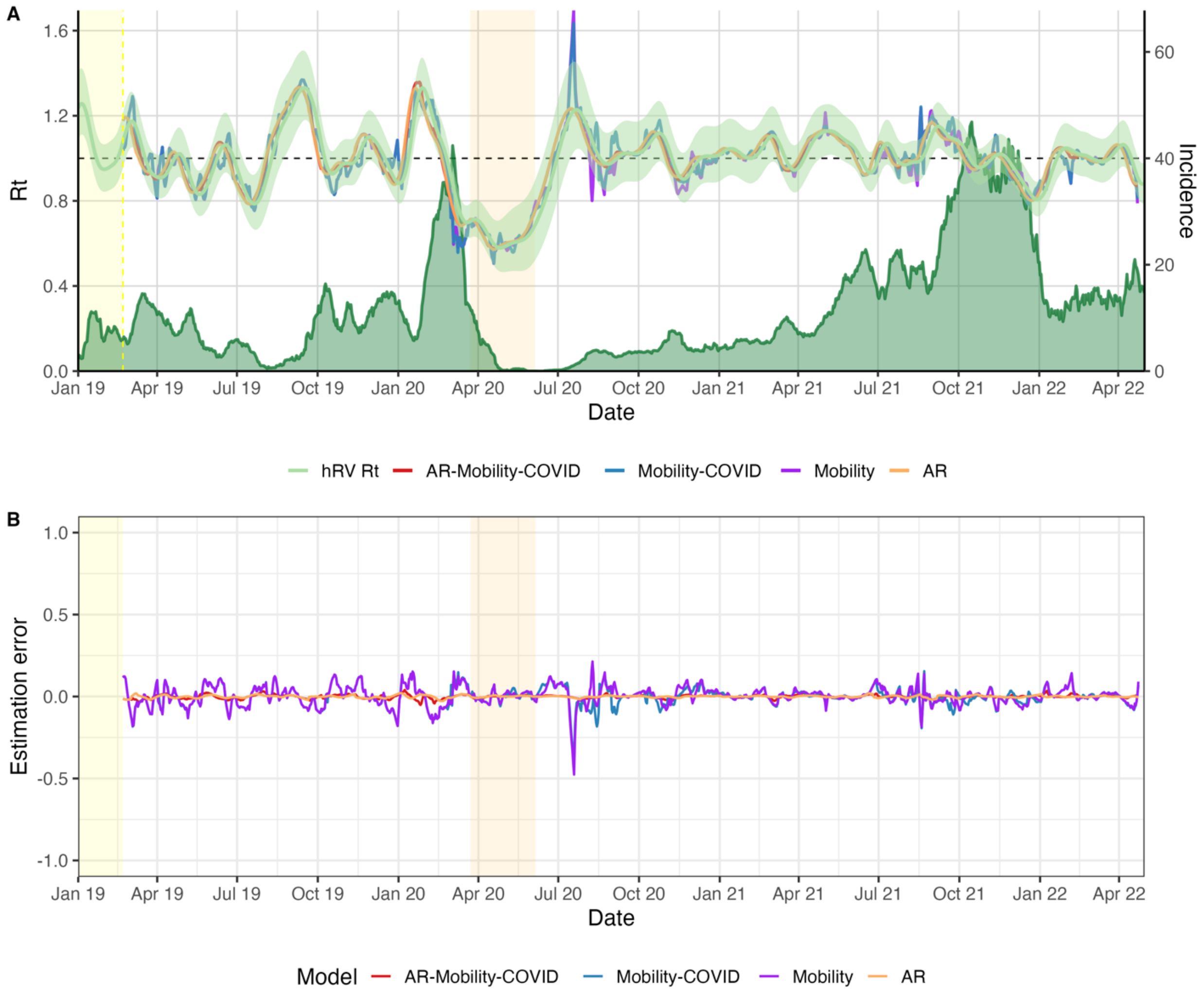


**Figure S19. Predictive model of daily human rhinovirus (hRV) transmission (time-varying effective reproduction number, Rt).** The yellow shaded area indicates the initial model training period, and the orange shaded area indicates Seattle's stay-at-home period. **A.** Estimated daily transmissibility (Rt) (left y-axis) from the full model with autoregressive (AR) terms, mobility terms, and SARS-CoV-2 interaction terms (red), contrasting with observed Rt (light green) and estimates from a model with only mobility and SARS-CoV-2 interaction terms (blue), a model with only mobility terms (purple), and a model with only AR terms (orange). Daily hRV incidence is shaded dark green (right y-axis). **B.** Model estimation error, defined as observed Rt minus predicted Rt.

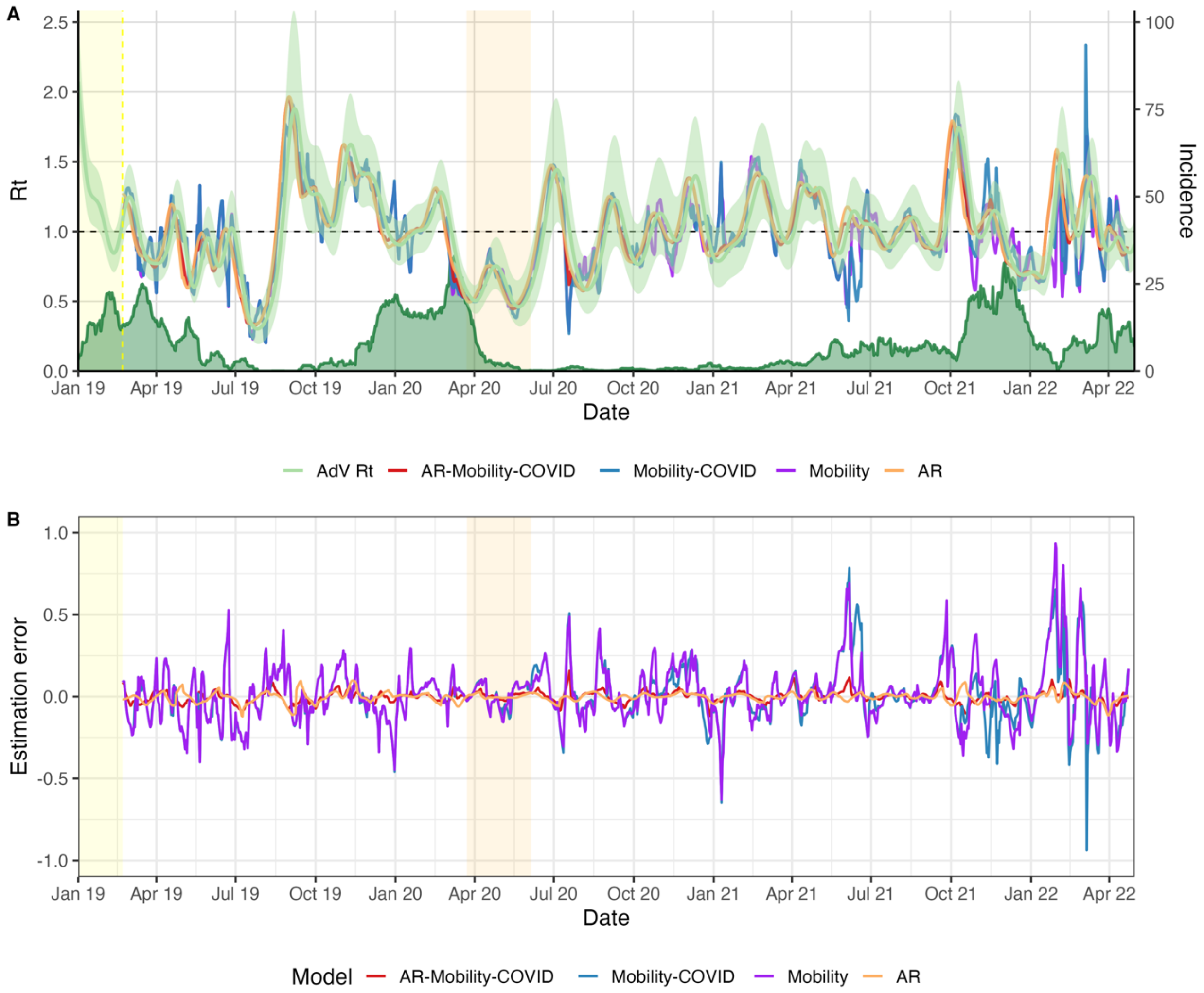


**Figure S20. Predictive model of daily adenovirus (AdV) transmission (time-varying effective reproduction number, Rt).** The yellow shaded area indicates the initial model training period, and the orange shaded area indicates Seattle's stay-at-home period. **A.** Estimated daily transmissibility (Rt) (left y-axis) from the full model with autoregressive (AR) terms, mobility terms, and SARS-CoV-2 interaction terms (red), contrasting with observed Rt (light green) and estimates from a model with only mobility and SARS-CoV-2 interaction terms (blue), a model with only mobility terms (purple), and a model with only AR terms (orange). Daily AdV incidence is shaded dark green (right y-axis). **B.** Model estimation error, defined as observed Rt minus predicted Rt.

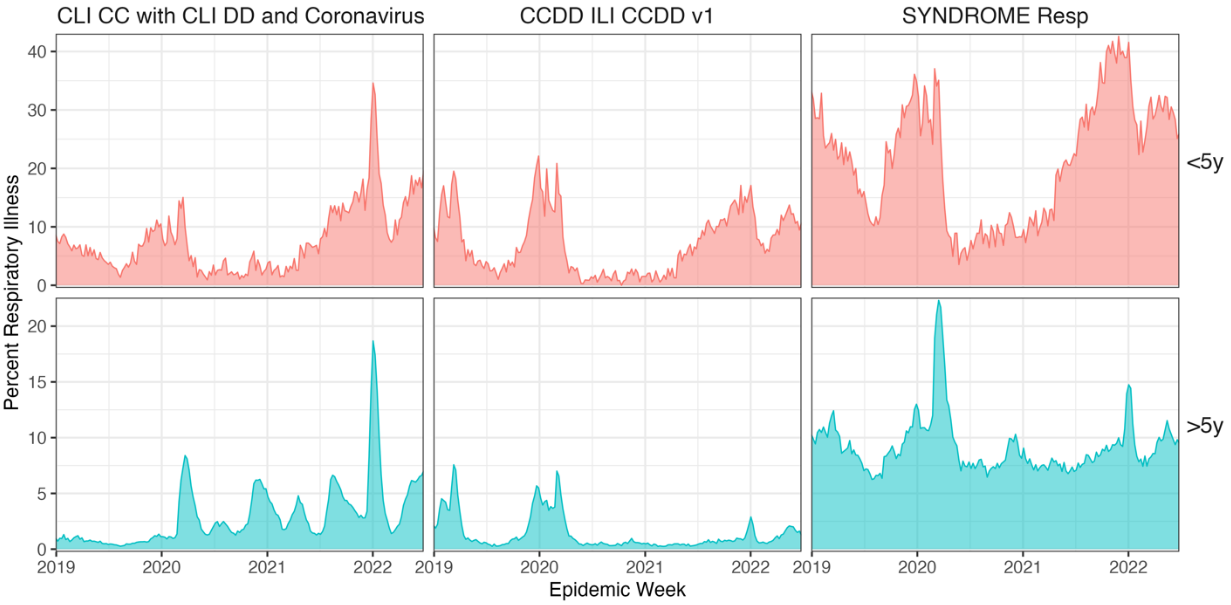


**Figure S21. The weekly proportion of emergency department visits coded as COVID-like illness (CLI), influenza-like illness (ILI), or broad respiratory illness among patients seeking care at emergency departments in King County, Washington.** Data are disaggregated by age group: < 5 (top) and ≥ 5 years of age (bottom). We applied a two-week rolling average reduce noise. Respiratory syndromic surveillance data for King County were obtained from the Rapid Health Information Network (RHINO) program at the Washington Department of Health. Syndrome criteria are defined by the Electronic Surveillance System for the Early Notification of Community-Based Epidemics (ESSENCE).

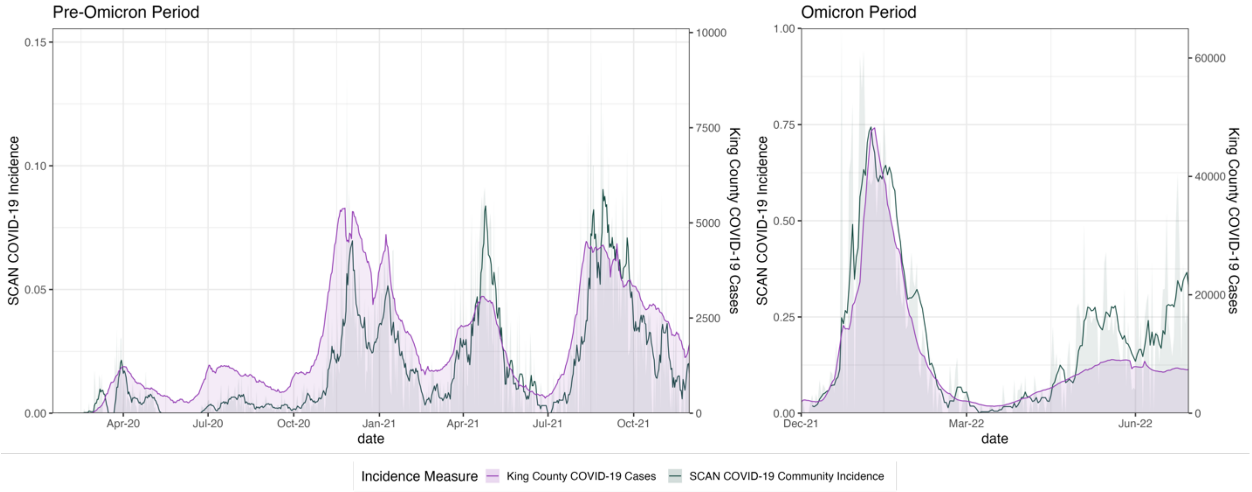


**Figure S22. Comparison of SCAN estimated daily COVID-19 incidence to daily King County COVID-19 cases.** Comparisons are split into two periods: pre-Omicron (before December 2021) and Omicron due to high case counts during the Omicron BA.1 wave in winter 2021-2022. We applied a two-week rolling average reduce noise.

### Supplementary Tables

**Table S1. OpenArray panel probe sets over time.** Green shaded areas indicate when a particular probe was in use.

| Pathogen type | Probe | V1<br>Start: 5-5-2019<br>End: 2-20-2020 | V2<br>Start: 2-21-2020<br>End: 5-1-2020 | V3<br>Start: 5-29-2020<br>End: 11-20-2020 | V4<br>Start: 11-23-2020 |
|---|---|---|---|---|---|
| **Influenza virus** | Flu_A_pan | | | | |
| | Flu_A_H1 | | | | |
| | Flu_A_H3 | | | | |
| | Flu_B_pan | | | | |
| | Flu_C | | | | |
| **Parainfluenza virus** | hPIV1_hPIV2_ | | | | |
| | hPIV3_hPIV4 | | | | |
| **Enterovirus** | EV_pan | | | | |
| | EV_D68 | | | | |
| **Rhinovirus** | RV_1of1 | | | | |
| | RV_1of2 | | | | |
| **Adenovirus** | AdV_1of1 | | | | |
| | AdV_1of2 | | | | |
| **Coronavirus** | CoV_HKU1_CoV_NL63 | | | | |
| | CoV_229E_CoV_OC43 | | | | |
| | hCoV_HKU1 | | | | |
| | hCoV_NL63 | | | | |
| | hCoV_229E | | | | |
| | hCoV_OC43 | | | | |
| | SARS_CoV-2_Orf1B | | | | |
| | SARS-CoV-2_S | | | | |
| **Respiratory Syncytial Virus** | RSVA | | | | |
| | RSVB | | | | |
| **Metapneumovirus** | hMPV | | | | |
| **Parechovirus** | hPeV | | | | |
| **Bocavirus** | hBoV | | | | |
| **Measles** | Measles | | | | |
| **Mumps** | Mumps | | | | |
| **Pneumoniae** | M. pneumoniae | | | | |
| | C. pneumoniae | | | | |
| | M. pneumo_C. pneumo | | | | |
| | S. pneumoniae | | | | |
| **Total unique pathogens** | | **26** | **26** | **26** | **24** |

**Table S2. Number of samples by recruitment type and site.**

| Recruitment Type | Site | Sample size |
|---|---|---|
| Clinic (Kiosk) | ChildrensHospitalSeattle | 944 |
| Clinic (Kiosk) | ChildrensHospitalSeattleOutpatientClinic | 246 |
| Clinic (Kiosk) | UWHallHealth | 196 |
| Clinic (Kiosk) | ChildrensHospitalBellevue | 94 |
| Clinic (Kiosk) | UWSeaMar | 88 |
| Clinic (Kiosk) | PioneerSquare | 58 |
| Clinic (Kiosk) | ChildrensSeaMar | 37 |
| Clinic (Flu VE Network) | Kaiser Permanente | 3604 |
| Community (swab-and-send) | SCAN | 42837 |
| Community (swab-and-send) | swabNSend | 2900 |
| Community (Residual) | RetrospectivePHSKC | 7934 |
| Community (Kiosk) | WestlakeMall | 392 |
| Community (Kiosk) | HarborviewLobby | 254 |
| Community (Kiosk) | UWSuzzalloLibrary | 177 |
| Community (Kiosk) | FredHutchLobby | 171 |
| Community (Kiosk) | HUB | 135 |
| Community (Kiosk) | Costco | 34 |
| Community (Kiosk) | CapitolHillLightRailStation | 32 |
| Community (Kiosk) | SeattleCenter | 23 |
| Community (Kiosk) | ColumbiaCenter | 18 |
| Community (Kiosk) | SeaTacDomestic | 12 |
| Community (Kiosk) | SeaTacInternational | 9 |
| Community (Kiosk) | PICAWA | 6 |
| Community (Kiosk) | KingStreetStation | 4 |
| Community (Kiosk) | WestlakeLightRailStation | 1 |
| Hospital (Residual) | RetrospectiveChildrensHospitalSeattle | 16025 |
| Hospital (Residual) | RetrospectiveHarborview | 2482 |
| Hospital (Residual) | RetrospectiveNorthwest | 1426 |
| Hospital (Residual) | RetrospectiveUWMedicalCenter | 707 |

**Table S3. Comparison of different models forecasting daily effective reproduction numbers (Rt) of human rhinovirus (hRV), adenovirus (AdV), and SARS-CoV-2.** The accuracy of 7-day ahead forecasts of hRV, AdV, and SARS-CoV-2 Rt were measured using the root-mean-squared error (RMSE), mean absolute error (MAE), mean absolute percentage error (MAPE), and Pearson correlation with observed Rt values.

### Time period: whole study period[1]

| 7-day ahead accuracy | Human rhinovirus (hRV) | | | | Adenovirus (AdV) | | | | SARS-CoV-2 | | | |
|---|---|---|---|---|---|---|---|---|---|---|---|---|
| | AR-Mobility-COVID Rt | Mobility-COVID Rt | Mobility | AR | AR-Mobility-COVID Rt | Mobility-COVID Rt | Mobility | AR | AR-Mobility-Rhino Rt | Mobility-Rhino Rt | Mobility | AR |
| **RMSE** | 0.011 | 0.058 | 0.059 | 0.007 | 0.035 | 0.173 | 0.174 | 0.029 | 0.031 | 0.129 | 0.151 | 0.029 |
| **MAE** | 0.008 | 0.042 | 0.042 | 0.005 | 0.026 | 0.125 | 0.125 | 0.021 | 0.02 | 0.086 | 0.099 | 0.018 |
| **MAPE** | 0.008 | 0.043 | 0.043 | 0.005 | 0.028 | 0.131 | 0.132 | 0.022 | 0.018 | 0.081 | 0.091 | 0.016 |
| **Correlation** | 1.0 | 0.93 | 0.93 | 1.0 | 0.99 | 0.83 | 0.82 | 0.99 | 0.99 | 0.87 | 0.83 | 1.0 |

[1] The hRV and AdV modeling periods include January 2019 – April 2022. The SARS-CoV-2 modeling period includes March 2020 – April 2022.

### Time period: stay-at-home orders and lifting of restrictions[2]

| 7-day ahead accuracy | Human rhinovirus (hRV) | | | | Adenovirus (AdV) | | | | SARS-CoV-2 | | | |
|---|---|---|---|---|---|---|---|---|---|---|---|---|
| | AR-Mobility-COVID Rt | Mobility-COVID Rt | Mobility | AR | AR-Mobility-COVID Rt | Mobility-COVID Rt | Mobility | AR | AR-Mobility-Rhino Rt | Mobility-Rhino Rt | Mobility | AR |
| **RMSE** | 0.006 | 0.055 | 0.049 | 0.005 | 0.021 | 0.097 | 0.091 | 0.019 | 0.031 | 0.133 | 0.122 | 0.028 |
| **MAE** | 0.005 | 0.042 | 0.037 | 0.004 | 0.017 | 0.077 | 0.072 | 0.014 | 0.02 | 0.086 | 0.085 | 0.019 |
| **MAPE** | 0.007 | 0.057 | 0.051 | 0.006 | 0.025 | 0.105 | 0.097 | 0.021 | 0.016 | 0.072 | 0.072 | 0.016 |
| **Correlation** | 1.0 | 0.93 | 0.94 | 1.0 | 1.0 | 0.94 | 0.95 | 1.0 | 0.99 | 0.93 | 0.94 | 0.99 |

[2] The hRV and AdV modeling periods include March – June 2020. The SARS-CoV-2 modeling period includes April – June 2020.

**Table S4. Data sources for adjusting the age distributions of pathogen presence/absence data.**

| | **Source** | **Description** | **Proportion < 5 years** | **Proportion ≥ 5 years** |
|---|---|---|---|---|
| **Community samples** | | | | |
| Influenza A/H3N2, A/H1N1, B | CDC FluView Interactive(*7*) | Age group distribution of influenza positive specimens reported by public health laboratories in WA state | Time varying (weekly) | Time varying (weekly) |
| AdV, hCoV, hMPV, hPIV, RSV, hRV | CDC FluView Interactive(*7*) | Age group distribution of influenza-like illness cases in WA state | Time varying (weekly) | Time varying (weekly) |
| SARS-CoV-2 | Washington Department of Health(*8*) | Age group distribution of COVID-19 positive specimens in King County, WA | Time varying (daily) | Time varying (daily) |
| **Hospital residuals** | | | | |
| Influenza A/H3N2, A/H1N1, B | Influenza Hospitalization Surveillance Network (FluSurv-NET)(*9*) | National age group distribution of laboratory-confirmed influenza-associated hospitalizations | Time varying (weekly) | Time varying (weekly) |
| RSV | Matias et al., 2017(*10*) | Age distribution of RSV detections among 186,155 positive patients hospitalized for ARI, cardiorespiratory disease, or sepsis, United States, 1997-2009 | 0.54 | 0.46 |
| hRV | El-Sahly et al., 2000(*11*) | Age distribution of hRV detections among 60 positive patients hospitalized for ARI, Houston, Texas, 1991-1995 | 0.4 | 0.6 |
| AdV | Akello et al., 2020(*12*) | Age distribution of AdV detections among 1302 positive patients referred to the Institute for Infectious Diseases for diagnostic testing, Bern, Switzerland, 1998-2017 | 0.57 | 0.43 |
| hCoV | Nickbakhsh et al., 2020(*13*) | Age distribution of hCoV detections among 2958 positive patients in secondary care, NHS Greater Glasgow and Clyde, Scotland, UK, 2005-2017 | 0.29 | 0.71 |
| hMPV | Barrera-Badillo et al., 2020(*14*) | Age distribution of hMPV detections among 331 positive patients hospitalized for SARI, Mexico, 2009-2018 | 0.575 | 0.425 |
| hPIV | Zhao et al., 2017(*15*) | Age distribution of hPIV detections among 17,717 positive patients in primary or secondary care, England and Wales, UK, 1998-2013 | 0.64 | 0.36 |
| SARS-CoV-2 | Washington Department of Health(*8*) | Age group distribution of laboratory confirmed COVID-19 hospitalizations in Washington state | Time varying (daily) | Time varying (daily) |

**Table S5. Pathogen-specific incubation periods and generation or serial intervals obtained from published literature.** Incubation periods and generation or serial intervals include the mean and standard deviation (SD) in days. The probability distribution family used to estimate each parameter is listed below the mean and SD.

| Pathogen | Incubation Period (days) | Generation or Serial Interval (days) | Source |
|---|---|---|---|
| SARS-CoV-2 | Mean = 6.3, SD = 3.6<br>Lognormal | Mean = 5.2, SD = 1.2<br>Gamma | Xin et al. 2021(*16*);<br>Ganyani et al. 2020(*17*) |
| hCoV * | Mean = 5.1, SD = 2.2<br>Lognormal | Mean = 5.2, SD = 1.2<br>Gamma | Spencer et al. 2022(*18*);<br>Ganyani et al. 2020(*17*) |
| Influenza | Mean = 1.9, SD = 1.22<br>Lognormal | Mean = 3.6, SD = 1.6<br>Weibull | Lessler et al. 2009(*19*);<br>Cowling et al. 2009 (*20*) |
| RSV ˆ | Mean = 4.5, SD = 0.9<br>Lognormal | Mean = 7.5, SD = 2.1<br>Gamma | Spencer et al. 2022(*18*);<br>Crowcroft et al. 2008(*21*) |
| hMPV ˆ† | Mean = 4.5, SD = 0.9<br>Lognormal | Mean = 5.2, SD = 1.5<br>Gamma | Spencer et al. 2022(*18*);<br>Matzuzaki et a. 2013(*22*) |
| hPIV ˆ‡ | Mean = 2.6, SD = 1.35<br>Lognormal | Mean = 7.5, SD = 2.1<br>Gamma | Lessler et al. 2009(*19*);<br>Crowcroft et al. 2008(*21*) |
| hRV ˆ | Mean = 2.36, SD = 1.1<br>Lognormal | Mean = 4.4, SD = 2.7<br>Gamma | Spencer et al. 2022(*18*);<br>Foy et al. 1988(*23*) |
| AdV ˆ | Mean = 5.6, SD = 1.26<br>Lognormal | Mean = 7.8, SD = 2.4<br>Gamma | Lessler et al. 2009(*19*);<br>Guo et al. 2020(*24*) |

* Generation interval for SARS-CoV-2.
ˆ Serial interval reanalyzed using time intervals of disease onset for infectors and infectees from the published study.
† Incubation period for RSV.
‡ Serial interval for RSV.

**Table S6. International Classification of Diseases, Tenth Revision (ICD-10) codes used to designate hospitalized patients as symptomatic for respiratory illness.**

| Condition | ICD-10 Code |
|---|---|
| Acute upper respiratory infections | J00-J06 |
| Influenza and pneumonia | J10-J18 |
| Other acute lower respiratory infections | J20-J22 |
| Other diseases of upper respiratory tract | J30-J39 |
| Chronic lower respiratory diseases | J40-J47 |
| Other respiratory diseases principally affecting the interstitium | J80-J84 |
| Suppurative and necrotic conditions of the lower respiratory tract | J85-J86 |
| Other diseases of the pleura | J90-J94 |
| Other diseases of the respiratory system | J96-J99 |
| COVID-19 | U07.1 |
| Otitis media | H65-H66 |
| Hemorrhage from respiratory passages | R04 |
| Cough | R05 |
| Abnormalities in breathing | R06 |
| Pain in throat and chest | R07 |
| Hypoxemia | R09.02 |
| Nasal congestion or postnasal drip | R09.8 |
| Fever, unspecified | R50.9 |
| Respiratory tuberculosis | A15 |
| Viral infection of unspecified site | B34 |
| Viral conjunctivitis | B30 |
| Streptococcus, Staphylococcus, and Enterococcus as the cause of diseases classified elsewhere | B95 |
| Adenovirus as the cause of diseases classified elsewhere | B97.0 |
| Enterovirus as the cause of diseases classified elsewhere | B97.1 |
| Coronavirus as the cause of diseases classified elsewhere | B97.2 |
| Respiratory syncytial virus as the cause of diseases classified elsewhere | B97.4 |
| Human metapneumovirus as the cause of diseases classified elsewhere | B97.81 |
